# Changing socioeconomic and geographic gradients in cardiovascular disease risk factors in India – Evidence from nationally representative household surveys

**DOI:** 10.1101/2022.11.11.22282234

**Authors:** Sarah Wetzel, Pascal Geldsetzer, Sneha Sarah Mani, Aashish Gupta, Kavita Singh, Mohammed K. Ali, Dorairaj Prabhakaran, Nikhil Tandon, Nikkil Sudharsanan

**Author notes:** <u>Corresponding author:</u> Name: Sarah Wetzel, MA, Address: Heidelberg Institute of Global Health, Faculty of Medicine, Heidelberg University, Im Neuenheimer Feld 130.3 69120 Heidelberg, Germany. Joint senior authors (Pascal Geldsetzer, Nikkil Sudharsanan). <u>Other Authors:</u>. Name: Pascal Geldsetzer, Address: Division of Primary Care and Population Health, Department of Medicine, Stanford University, 3180 Porter Drive, Palo Alto, CA 94304, USA. Name: Sneha Sarah Mani, Address: Graduate Group in Demography, University of Pennsylvania, 207, 3718 Locust Walk, Philadelphia, PA 19104, USA. Name: Aashish Gupta, Address: Harvard University, 9 Bow Street, Cambridge, MA 02138, USA. Name: Kavita Singh, Address: Heidelberg Institute of Global Health, Faculty of Medicine, Heidelberg University, Im Neuenheimer Feld 130.3, 69120 Heidelberg. Name: Mohammed K. Ali, Address: Emory University, 1516 Clifton Rd NE, R755, Atlanta, GA 30322, USA. Name: Dorairaj Prabhakaran, Address: Centre for Chronic Disease Control, C1/52, Safdarjung Development Area New Delhi 110016, India. Name: Nikhil Tandon, Address: Department of Endocrinology and Metabolism, All India Institute of Medical Sciences Ansari Nagar, New Delhi 110029, India. Name: Nikkil Sudharsanan, Address: Professorship of Behavioral Science for Disease Prevention and Health Care, Technical University of Munich Georg-Brauchle-Ring 60, Room L420, 80992 Munich, Germany.

## Abstract

**Background:** Cardiovascular diseases (CVDs) are the leading cause of death in most low- and middle-income countries (LMICs). CVDs and their metabolic risk factors have historically been concentrated among urban residents with higher socioeconomic status (SES) in LMICs such as India. However, as India develops, it is unclear whether these socioeconomic and geographic gradients will persist or change. Understanding these social dynamics in CVD risk is essential for mitigating the rising burden of CVDs and to reach those with the greatest needs.

**Methods:** Using nationally representative data with biomarker measurements from the fourth (2015-16) and fifth (2019-21) Indian National Family and Health Surveys, we investigated trends in the prevalence of four CVD risk factors: tobacco consumption (self-reported, any type), unhealthy weight 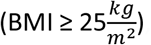, idiabetes (random plasma glucose concentration ≥200mg/dL or self-reported diabetes), and hypertension (one of: average systolic blood pressure ≥ 140mmHg, average diastolic blood pressure ≥ 90mmHg, self-reported past diagnosis, or self-reported current antihypertensive medication use) among adults aged 15-49 years. We first described changes at the national level and then trends stratified by place of residence (urban versus rural), geographic region (northern, northeastern, central, eastern, western, southern), regional level of development (Empowered Action Group member state or not), and two measures of socioeconomic status: level of education (no education, primary incomplete, primary complete, secondary incomplete, secondary complete, higher) and wealth (quintiles).

**Findings:** Unhealthy weight increased among all social and geographic groups but both the absolute and the relative changes were substantially higher among people with low SES (as measured by education or wealth) and in rural areas. For diabetes and hypertension, the prevalence increased for those from disadvantaged groups while staying constant or even decreasing among the wealthier and more educated. In contrast, tobacco consumption declined for all social and geographic groups.

**Interpretation:** In 2015-16, CVD risk factors were higher among more advantaged subpopulations in India. However, between 2015-16 and 2019-21, the prevalence of these risk factors grew more rapidly for less wealthy and less educated subpopulations and those living in rural areas. These trends have resulted in CVD risk becoming far more widespread throughout the population; CVD can no longer be characterized as a wealthy urban phenomenon.

**Funding:** This work was supported by the Alexander von Humboldt Foundation [NS]; the Stanford Diabetes Research Center [PG], and the Chan Zuckerberg Biohub [PG].

**Research in Context:** *Evidence before this study:* We searched PubMed for work published between Jan 1, 1990, and Sep 23, 2022, with variations of the search terms “reversal hypothesis”, “social gradient”, “socioeconomic gradient”, “social difference”, “socioeconomic difference”, “socioeconomic status”, “change”, “trend”, “cardiovascular disease”, “cardiovascular risk factor”, “diabetes”, “hypertension”, “overweight”, “obesity”, “smoking”, “tobacco”, “low-income”, “lower-middle-income”, and “India” in the title or abstract. Existing studies on changes in the socioeconomic patterning of cardiovascular disease (CVD) risk factors mostly compared the size of social gradients in obesity cross-sectionally between countries at different levels of national income or development. These studies generally found higher obesity among higher socioeconomic status (SES) populations but opposite gradients for countries at higher levels of development. However, because these studies use cross-sectional comparisons, whether these patterns reflect the influence of development or other contextual factors associated with countries at different levels of development is unclear. Both for obesity and for other CVD risk factors, we found few nationally-representative studies that traced how gradients within countries changed over time as they developed. Importantly, most of these studies did not focus on India and those that did only considered single risk factors such as obesity using older data. Existing nationally representative research on India is thus predominantly cross-sectional and has only focused on the size of social gradients at single points in time. This work generally finds higher levels of unhealthy weight, diabetes, and hypertension among wealthy and educated subpopulations but lower levels of tobacco consumption. Subnational and smaller non-representative studies from India that traced changes in CVD risk factors over time provide some evidence of reversing or flattening gradients but it is unclear whether these patterns hold at regional and national levels.

*Added value of this study:* Using data from the 2015-16 and 2019-2021 National Family and Health Surveys, we provide some of the first nationally representative evidence for trends in the prevalence of major CVD risk factors in India and how these trends have changed across social and geographic groups. Both absolute and relative increases in prevalence were far more pronounced among relatively poor and less educated populations. As a result of these trends, most CVD risk factors became more equal across population groups, or in some cases, became higher among the less compared to more educated.

*Implications of all the available evidence:* Indians with lower SES – as measured by wealth and education – experienced considerable adverse trends in unhealthy weight, diabetes, and hypertension between 2015-16 and 2019-21. Our results reveal that CVDs can no longer be considered a problem of the affluent parts of society and suggest that CVD prevention efforts that reach less advantaged subpopulations are urgently needed.

## Introduction

Cardiovascular diseases (CVDs) are the leading worldwide causes of death, accounting for 32% of all deaths globally. ^1,2^ The risk of CVD can be substantially reduced by addressing key behavioral and metabolic risk factors, particularly tobacco consumption, hypertension, diabetes, and unhealthy weight. ^3,4^ For this reason, monitoring the prevalence of these key risk factors is essential to identify populations at high risk and develop targeted public health interventions.

Previous studies from India and from other low- and middle-income countries (LMICs) generally find a higher prevalence of hypertension, diabetes, and unhealthy weight among people with higher compared to those with lower socioeconomic status (SES). ^5–8^ For example, based on data from the 2015-16 Indian National Family and Health Survey (NFHS), the prevalence of obesity was 18·6% among individuals in the highest household wealth quintile compared to just 1·7% among those in the lowest wealth quintile. Similarly, the prevalence of hypertension was 16·1% in urban compared to 13·4% in rural areas. ^7^ Such evidence of CVD risk as an urban and upper-class phenomenon has resulted in calls to prioritize health conditions like maternal and child health or infectious diseases instead of allocating public resources to CVD care. ^9^

In contrast to India and other LMICs, high-income countries and several upper-middle-income countries are typically characterized by a higher prevalence of CVD risk factors among disadvantaged subpopulations. ^10–12^ This stark difference has led researchers to hypothesize that while CVD risk is initially more concentrated among higher SES populations, as countries develop, this pattern will reverse and CVDs will become more pronounced among poorer subpopulations. ^10,13,14^ If true, CVD risk factors in India may also be changing and becoming more widespread throughout the population. Indeed, recent studies have found that some CVD risk factors like hypertension are equally high across all wealth groups in similar LMICs like Indonesia, ^14–17^ and subnational analyses within India also find first signs of reversal in more developed parts of the country. ^5,6,18^

It is currently unknown how SES differences in CVD risk factors (known as SES gradients) have changed over time in India. This gap is likely because repeated nationally representative data has not been available so far for many major CVD risk factors. ^19^ We address this key research gap by taking advantage of recently released nationally representative survey data with biomarker measurements. We begin by providing comprehensive evidence on changes in the prevalence of major behavioral and biological CVD risk factors. After showing trends at the national level, we measure changes separately across socioeconomic and geographic groups. Finally, we examine the implications of these trends for SES and geographic gradients and assess whether India is undergoing a shift of CVD risk towards more disadvantaged segments of the population.

## Methods

### Data

We used data from two consecutive nationally representative cross-sectional household surveys from India, the NFHS-4 and the NFHS-5. Both surveys used a complex two-stage stratified sampling design and a nearly identical sampling strategy. Commissioned by the Ministry of Health and Family Welfare, Government of India and with technical assistance from the Demographic and Health Surveys Program, the International Institute for Population Sciences (IIPS) conducted the two surveys from 2015 to 2016 (NFHS-4) and from 2019 to 2021 (NFHS-5). First, the IIPS selected primary sampling units (PSUs) within strata applying systematic random sampling with probability proportional to size. Villages and Census Enumeration Blocks served as PSUs in rural and urban areas, respectively. Afterwards, the institute chose 22 households from each PSU or – in PSUs with more than 300 households – from 2 out of three segments of a PSU (again through systematic random sampling). ^19,20^

Both the NFHS-4 and the NFHS-5 included questions on self-reported tobacco use (smoking and smokeless variants), current diabetes status (based on the question “Do you currently have diabetes?”), clinical history of hypertension (based on the questions “Were you told on two or more different occasions by a doctor or other health professional that you had hypertension or high blood pressure?” (NFHS-4) and “Has a doctor, nurse or auxiliary nurse midwife told you two or more times that you have hypertension or high blood pressure?” (NFHS-5)), and current intake of prescribed antihypertensive medication (based on the question “Are you now taking prescribed medication to lower your blood pressure?”). Furthermore, trained enumerators also collected anthropometric and biomarker measurements, including height, weight, random blood glucose, and three blood pressure readings with 5-minute intervals between measurements. In the NFHS-5, adults aged 15 years and older were eligible for glucose tests, blood pressure measurements, and asked about hypertension diagnoses or treatments while all other mentioned CVD-related items (diabetes status, tobacco use, height, and weight) in the NFHS-5 and all CVD items in the NFHS-4 only covered men aged 15-54 in a representative subsample of 15% of households (state module) and women aged 15-49 years.

The methods to measure biomarkers were consistent across survey rounds and the devices nearly equivalent. Weight, height, glucose, and blood pressure measurements were measured using the Seca 874 digital scale, the Seca 213 stadiometer, the FreeStyle Optium H glucometer (NFHS-4) or the Accu-Chek Performa glucometer (NFHS-5), and an Omron Blood Pressure Monitor, respectively.

### Main outcomes and variable definitions

We analyzed trends in four CVD risk factors: tobacco consumption, unhealthy weight, hypertension, and diabetes. We measured tobacco use as a binary indicator for self-reported tobacco consumption. Due to the widespread use of smokeless tobacco in India, ^21–23^ we considered not only self-reported cigarette smoking but also use of any other kind of tobacco.

We measured unhealthy weight using a binary indicator for overweight or obesity. For our main analysis, we classified participants with a BMI of at least 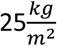 as having overweight or obesity. In the appendix, we also present results for trends in unhealthy weight using an indicator for obesity only (defined as 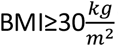) (**Figures S11-S13 and S37-S39** in the appendix).

We measured diabetes with a binary indicator for self-reported diabetes status or a random plasma glucose concentration 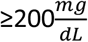. Our diabetes definition does not consider antidiabetic medication, since the NFHS-4 survey instruments did not include this question. The glucometers used by the NFHSs provided capillary blood glucose measurements which we multiplied by 1·11 to obtain plasma equivalents. ^19,20,24^

We measured hypertension with a binary indicator if at least one out of four conditions was satisfied: the respondent reported a past diagnosis by a clinician, reported being on an antihypertensive medication, had an average systolic blood pressure greater than or equal to 140mmHg, or had an average diastolic blood pressure greater than or equal to 90mmHg. As the first blood pressure measurement generally tends to be higher and might therefore overestimate the prevalence of hypertension, ^25,26^ we calculated average systolic and diastolic blood pressure based on the second and third reading. If one of these two readings was missing, we replaced it by the first. We excluded participants with only a single reading of systolic or diastolic blood pressure. We also excluded individuals with implausible biomarker values: systolic blood pressure below 70 mmHg or above 240 mmHg, diastolic blood pressure below 40 mmHg or above 130 mmHg, or random blood glucose below 40 mg/dL. If any of the variables needed to define an indicator was not available, we set the respective indicator to missing.

### Socioeconomic and geographic subgroups

As part of our analysis, we stratified results by sex (measured by a binary variable for male versus female), age (measured in 5-year age groups), level of education (classified as: no education, primary incomplete, primary complete, secondary incomplete, secondary complete, or higher), household wealth (measured in quintiles of a continuous index based on household asset ownership, housing materials, and water and sanitation access^27,28^), place of residence (measured by a binary variable for urban versus rural areas), geographic region (grouping northern, northeastern, central, eastern, western, and southern administrative divisions as defined by the NFHS^20^), and regional level of development measured by a binary variable for households living in one of the eight Empowered Action Group (EAG) states (Bihar, Chhattisgarh, Jharkhand, Madhya Pradesh, Orissa, Rajasthan, Uttarakhand, and Uttar Pradesh). In 2001, the Government of India constituted this EAG with the aim of fostering development in India’s less developed states. ^29^ Except for the location-based indicators, all these variables were based on household or respondent self-reports.

### Statistical methods

We restricted our analysis to individuals aged 15-49 who, regardless of sex and survey round, were eligible for biomarker measurements and asked about their health behavior/status. Furthermore, we excluded pregnant women and participants who replied with “Don’t know” when asked about their level of education. To arrive at the final sample for the analysis of each outcome, we dropped observations with missing values in the respective dependent variable (i.e., the sample sizes differed depending on the outcome).

After describing the unweighted samples by survey year, we first report year-specific prevalence levels, absolute changes, and changes in percentage terms relative to the NFHS-4 at the national level. We estimated these changes over time using log binomial regression models for each of the CVD risk factors as the dependent variables and a binary indicator for the NFHS-5 as our main independent variable which captures the overall change in each CVD risk factor over time. We use the estimated coefficient on the survey indicator to present results for the change in CVD risk factor on both the absolute and relative scales.

We then measured changes in the risk factors across socioeconomic and geographic groups to identify disproportionately affected subpopulations and determine how gradients have changed over time. To do this, we estimated similar regression models as for the national level additionally including one of the grouping variables (to capture the baseline social gradients in the NFHS-4) and an interaction term between the survey year dummy and each social and geographic grouping variable (to capture how the gradients have changed over time). We only included one grouping variable in the regression model at a time.

All steps of our analysis factor in the complex survey design, including the survey weights. However, we performed two adjustments to the original NFHS weights: given the subsampling of men as described above, all outcomes except for the hypertension indicator in the NFHS-5 were only collected for a subset of men aged 15-49 but for all women in this age group. We rescaled the survey weights of men in the datasets for the analysis of diabetes, smoking, and unhealthy weight and the survey weights of male observations from the NFHS-4 in the hypertension dataset to ensure an equal representation of men and women. Secondly, we applied direct age standardization to the survey weights (using the age distribution of the WHO standard population) ^30^ such that all our estimates of CVD trends are net of demographic changes as Asian countries are faced with substantial aging and CVD risk factors are positively correlated with age. ^31–33^ In the appendix (**Table S2 and Figures 2-31**), we also show unadjusted changes based on the Indian age distribution in the respective survey year according to the United Nations World Population Prospects from 2019 (UNWPP). ^34^ We used R version 4·1·0 and Stata 17·0 for our analysis.

## Results

### Sample characteristics

Taken together, the NFHS-4 and the NFHS-5 collected data from 2,350,441 individuals aged 15-49 (NFHS-4: 866,304, NFHS-5: 1,484,137). After excluding pregnant women, people with unknown level of education, and participants with missing outcome variable, the final samples for the analysis of smoking, unhealthy weight, hypertension, and diabetes were 1,558,135 (NFHS-4: 769,373, NFHS-5: 788,762), 1,539,701 (NFHS-4: 768,881, NFHS-5: 770,820), 1,993,780 (NFHS-4: 757,047, NFHS-5: 1,236,733), and 1,481,174 (NFHS-4: 740,078, NFHS-5: 741,096) individuals, respectively. A flowchart showing the sampling procedure is available in the appendix (**Figure S1**).

The unweighted social and demographic characteristics were similar across the different outcome samples and survey rounds (**Table 1**). Around 88% of all the samples consisted of female respondents, except for the NFHS-5 hypertension sample (47% female), and between 50-52% of the samples were above the age of 30. With respect to education, between 17-25% of the samples had no formal education and between 12-16% completed more than secondary education. The share of respondents from urban households ranged from 25-30%. We present the weighted descriptive statistics in **Table S1** in the appendix.

**Table 1.** **Unweighted descriptive characteristics of the study samples, National Family Health Survey 4 (2015-16) and 5 (2019-21), India**

|  | Tobacco sample |  | Overweight sample |  | Diabetes sample |  | Hypertension sample |  |
| --- | --- | --- | --- | --- | --- | --- | --- | --- |
|  | NFHS-4<br>N = 769,373 | NFHS-5<br>N = 788,762 | NFHS-4<br>N = 768,881 | NFHS-5<br>N = 770,820 | NFHS-4<br>N = 740,078 | NFHS-5<br>N = 741,096 | NFHS-4<br>N = 757,047 | NFHS-5<br>N = 1,236,733 |
| **Sex** |  |  |  |  |  |  |  |  |
| Male | 103,297 (13%) | 93,231 (12%) | 102,788 (13%) | 89,676 (12%) | 98,673 (13%) | 85,715 (12%) | 101,049 (13%) | 577,017 (47%) |
| Female | 666,076 (87%) | 695,531 (88%) | 666,093 (87%) | 681,144 (88%) | 641,405 (87%) | 655,381 (88%) | 655,998 (87%) | 659,716 (53%) |
| **Age group** |  |  |  |  |  |  |  |  |
| 15-19 years | 140,138 (18%) | 136,151 (17%) | 139,926 (18%) | 131,249 (17%) | 134,750 (18%) | 127,141 (17%) | 137,264 (18%) | 211,984 (17%) |
| 20-24 years | 125,648 (16%) | 121,909 (15%) | 126,383 (16%) | 119,284 (15%) | 121,160 (16%) | 114,828 (15%) | 124,393 (16%) | 190,099 (15%) |
| 25-29 years | 121,164 (16%) | 123,519 (16%) | 121,450 (16%) | 121,223 (16%) | 116,698 (16%) | 116,886 (16%) | 119,573 (16%) | 192,317 (16%) |
| 30-34 years | 107,797 (14%) | 110,709 (14%) | 107,524 (14%) | 108,540 (14%) | 103,880 (14%) | 104,630 (14%) | 105,996 (14%) | 174,045 (14%) |
| 35-39 years | 102,906 (13%) | 109,703 (14%) | 102,568 (13%) | 107,445 (14%) | 98,963 (13%) | 103,173 (14%) | 101,095 (13%) | 172,145 (14%) |
| 40-44 years | 88,146 (11%) | 91,965 (12%) | 87,806 (11%) | 90,142 (12%) | 84,641 (11%) | 85,993 (12%) | 86,627 (11%) | 145,708 (12%) |
| 45-49 years | 83,574 (11%) | 94,806 (12%) | 83,224 (11%) | 92,937 (12%) | 79,986 (11%) | 88,445 (12%) | 82,099 (11%) | 150,435 (12%) |
| **Level of education** |  |  |  |  |  |  |  |  |
| No education | 192,447 (25%) | 163,166 (21%) | 192,895 (25%) | 160,822 (21%) | 186,123 (25%) | 153,789 (21%) | 190,622 (25%) | 205,670 (17%) |
| Incomplete primary | 49,529 (6.4%) | 45,449 (5.8%) | 49,555 (6.4%) | 44,845 (5.8%) | 47,387 (6.4%) | 42,804 (5.8%) | 48,732 (6.4%) | 69,730 (5.6%) |
| Complete primary | 52,918 (6.9%) | 52,543 (6.7%) | 53,205 (6.9%) | 51,560 (6.7%) | 51,140 (6.9%) | 49,445 (6.7%) | 52,595 (6.9%) | 79,585 (6.4%) |
| Incomplete secondary | 312,489 (41%) | 331,035 (42%) | 312,251 (41%) | 323,582 (42%) | 300,305 (41%) | 311,635 (42%) | 306,889 (41%) | 545,846 (44%) |
| Complete secondary | 69,671 (9.1%) | 83,246 (11%) | 69,620 (9.1%) | 81,009 (11%) | 67,082 (9.1%) | 78,251 (11%) | 68,385 (9.0%) | 142,878 (12%) |
| Higher than secondary | 92,319 (12%) | 113,323 (14%) | 91,355 (12%) | 109,002 (14%) | 88,041 (12%) | 105,172 (14%) | 89,824 (12%) | 193,024 (16%) |
| **Wealth quintile** |  |  |  |  |  |  |  |  |
| Poorest | 142,309 (18%) | 161,000 (20%) | 143,283 (19%) | 158,867 (21%) | 137,492 (19%) | 151,741 (20%) | 141,458 (19%) | 254,419 (21%) |
| Poorer | 163,112 (21%) | 174,522 (22%) | 163,967 (21%) | 172,049 (22%) | 157,087 (21%) | 165,079 (22%) | 161,436 (21%) | 274,795 (22%) |
| Middle | 162,850 (21%) | 165,645 (21%) | 163,093 (21%) | 162,821 (21%) | 156,870 (21%) | 156,860 (21%) | 160,251 (21%) | 262,149 (21%) |
| Richer | 154,148 (20%) | 152,987 (19%) | 153,554 (20%) | 149,184 (19%) | 148,197 (20%) | 144,074 (19%) | 151,046 (20%) | 239,907 (19%) |
| Richest | 146,954 (19%) | 134,608 (17%) | 144,984 (19%) | 127,899 (17%) | 140,432 (19%) | 123,342 (17%) | 142,856 (19%) | 205,463 (17%) |
| **Place of residence** |  |  |  |  |  |  |  |  |
| Urban | 229,285 (30%) | 198,099 (25%) | 226,286 (29%) | 189,467 (25%) | 218,123 (29%) | 182,202 (25%) | 222,783 (29%) | 306,503 (25%) |
| Rural | 540,088 (70%) | 590,663 (75%) | 542,595 (71%) | 581,353 (75%) | 521,955 (71%) | 558,894 (75%) | 534,264 (71%) | 930,230 (75%) |

### Trends at the national level

From 2015-16 to 2019-21, the largest absolute increase in prevalence occurred for unhealthy weight (overweight or obesity: 3·4 percentage points, 95% CI: 3·0, 3·8; obesity: 1·2 percentage points, 95% CI 1·0, 1·3) followed by diabetes (0·3 percentage points, 95% CI 0·2, 0·5) (**Table 2**). While the absolute changes appear small, these increasing trends corresponded to pronounced relative changes when compared to the baseline NFHS-4 values: overweight or obesity increased by 17% (95% CI 14·7, 19·3) from a baseline prevalence of 20% (95% CI 19·7; 20·3), obesity by 28·4% (95% CI 23·7, 33·2) from a baseline prevalence of 4·1% (95% CI 4·0, 4·2), and diabetes by 9·5% (95% CI 4·4, 14·7) from a baseline prevalence of 3·3% (95% CI 3·2, 3·4). We found no evidence for a meaningful national-level change in hypertension (−0·2 percentage points, 95% CI -0·5, 0·2).

**Table 2.** **Levels, absolute and relative changes in 4 CVD risk factors at the national level in India based on the National Family Health Survey 4 (2015-16) and 5 (2019-21)**

| Outcome | NFHS-4 | NFHS-5 | Absolute change<br>(percentage points) | Relative change (%) |
| --- | --- | --- | --- | --- |
| Tobacco consumption | 26.5 [26.1; 26.9] | 21.1 [20.7; 21.5] | -5.4 [-6; -4.8] | -20.4 [-22.3; -18.4] |
| Overweight or obesity | 20 [19.7; 20.3] | 23.4 [23.1; 23.6] | 3.4 [3; 3.8] | 17 [14.7; 19.3] |
| Diabetes | 3.3 [3.2; 3.4] | 3.6 [3.5; 3.7] | 0.3 [0.1; 0.5] | 9.5 [4.4; 14.7] |
| Hypertension | 17.8 [17.5; 18.1] | 17.6 [17.5; 17.8] | -0.2 [-0.5; 0.2] | -0.9 [-2.8; 1.1] |

Over the period, tobacco consumption decreased by 5·4 percentage points nationally (95% CI -6·0, - 4·8). Most of this decline was attributable to a lower consumption of tobacco variants other than cigarettes (**Table S3**). In relative terms, this change corresponded to a 20·4% (95% CI -22·3, -18·4] decline in the share of the population that uses tobacco.

### Changing socioeconomic gradients

In 2015-16, the prevalence of both overweight and obesity was higher among wealthier and educated NFHS-4 participants (**Figure S36 and Figure S37** in the appendix). However, the magnitude of the change in overweight and obesity between 2015-16 and 2019-21 was substantially higher among people with low SES compared to people with high SES (**Figures 1 and 2**): Overweight increased by 4·5 percentage points (95% CI 4·0, 5·1) among Indians without education compared to just 2·1 percentage points (95% CI 0·8, 3·4) among the most educated. This corresponded to a 29·5% (95% CI 25·7, 33·6) increase among the uneducated compared to a just 7·6% (95% CI 2·8, 12·6) increase among those with the highest level of education. Similarly, the poorest 20% were faced with a stark rise of 4·3 percentage points (95% CI 3·9, 4·7) in overweight while the richest 20% of the population experienced a more modest change of 2·9 percentage points (95% CI 1·8, 4·1). In relative terms, these upward trends were equivalent to increases of 78·7% (95% CI 68·9, 89·0) in the first and 8·3% (95% CI 4·9, 11·9) in the fifth quintile.

**Figure 1.**
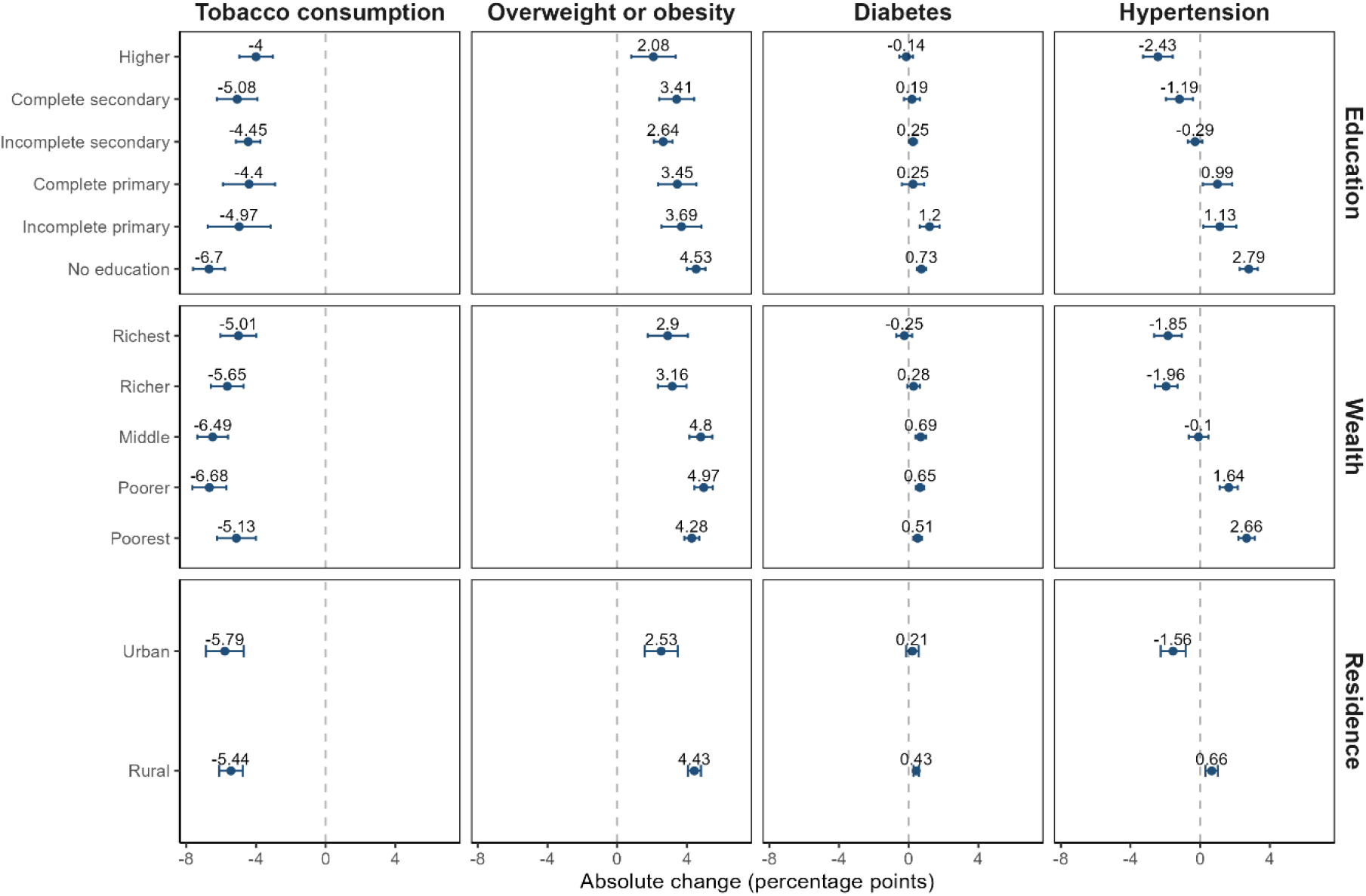
**Age-adjusted absolute change in each CVD risk factor among adults aged 15-49 by wealth quintile, level of education, and place of residence, National Family Health Survey 4 (2015-16) and 5 (2019-21), India**

**Figure 2.**
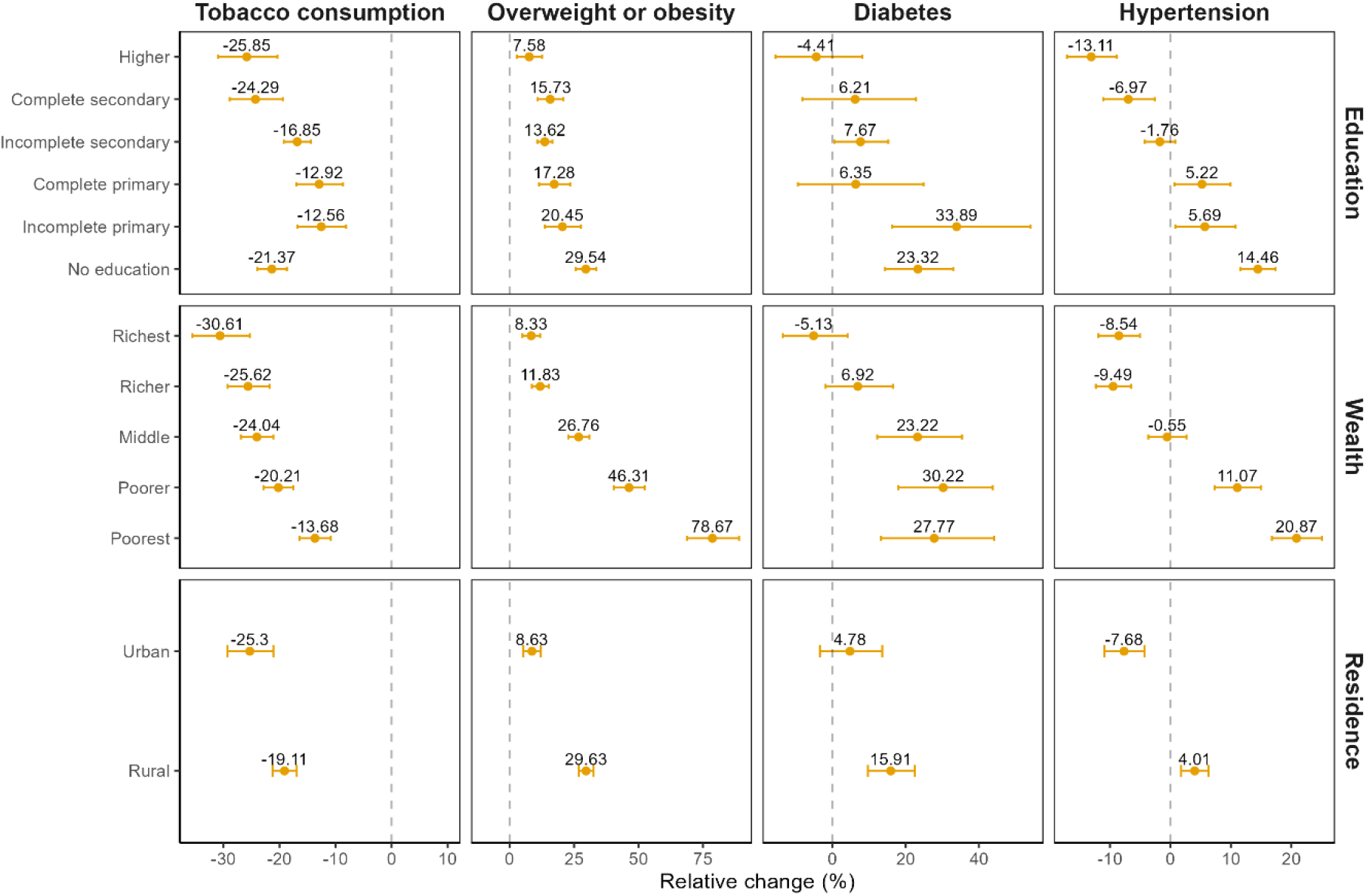
**Age-adjusted relative change in each CVD risk factor among adults aged 15-49 by wealth quintile, level of education, and place of residence, National Family Health Survey 4 (2015-16) and 5 (2019-21), India**

For diabetes, the size of the SES gradient in 2015-16 varied across the two measures of SES (**Figure S40** in the appendix). Across wealth, the prevalence of diabetes was 1·8% (95% CI 1·7, 2·0) among those in the poorest wealth quintile compared to 5·0% (95% CI 4·7, 5·3) in the highest wealth quintile. In contrast, the prevalence of diabetes in 2015-16 was similar across all education groups. Despite these baseline differences, we find that the increase in the prevalence of diabetes between 2015-16 and 2019-21 was larger for disadvantaged groups across both measures of SES (**Figures 1 and 2**). The prevalence of diabetes increased by 0·5 percentage points (95% CI 0·3, 0·8; relative increase of 27·8%) for those in the bottom wealth quintile compared to a 0·3 percentage points decrease (95% CI -0-7, 0·2; relative decrease of 5·1%) for those in the top wealth quintile. Among those without any education, diabetes rose by 0·7 percentage points (95% CI 0·5, 1·0; relative increase of 23·3%), whereas prevalence among people with more than secondary education remained virtually unchanged (−0·1 percentage points, 95% CI -0·5, 0·3). These changes led to a substantial flattening in the wealth gradients and a complete reversal in education gradients for diabetes between 2015-16 and 2019-21.

Like diabetes, we find that individuals with lower levels of education and wealth experienced a much larger change in the prevalence of hypertension between 2015-16 and 2019-21 (**Figures 1 and 2**). Hypertension increased by 2·8 percentage points (95% CI 2·3, 3·3; relative change of 14·5%) for those without education but decreased by 2·4 percentage points (95%CI -3·3, -1·6; relative decrease of - 13·1%) for those with the highest level of education. Likewise, hypertension rose by 2·7 percentage points (95% CI 2·2, 3·1; relative increase of 20·9%) in the poorest wealth quintile while it declined by 1·9 percentage points (95% CI -2·6, -1·1; 8·5% relative decline) in the top wealth quintile. Due to these changes over time, SES gradients in hypertension measured by education have fully reversed and the prevalence of hypertension is now higher among those with lower compared to higher SES. As of 2019-21, the prevalence of hypertension is 16·1%, (95% CI 15·8, 16·4) among the most educated compared to 22·1%, (95% CI 21·8, 22·4) among those without education.

Both the baseline gradients and trends over time in tobacco use differed substantially from the other CVD risk factors. In 2015-16, tobacco use was greater among those in the lowest (37·5%, 95% CI 36·7, 38·3) compared to the highest (16·4%, 95% CI 15·5, 17·2) wealth quintiles, and, among those with no education (24·7%, 95% CI 24·0, 25·3) compared to those with more than secondary education (11·5%, 95% CI 10·9, 12·1) (**Figure S32** in the appendix). Between 2015-16 and 2019-21, the absolute decline was greater for those with lower SES; however, because of the higher initial prevalence, this absolute decline corresponded to a smaller relative change compared to those with higher SES (**Figures 1 and 2**). For example, tobacco use declined by 6·7 percentage points (95% CI -7·6, -5·8) for those with no education compared to 4·0 percentage points (95% CI -5·0, -3·0) among those with the highest level of education. However, this corresponded to a 21·4% decrease (95% CI -24·0, -18·7) for the uneducated compared to a 25·9% decrease (95% CI -30·9, -20·4) for the most educated group.

These shifts in the burden of unhealthy weight, hypertension, and diabetes to people with low SES affected both sexes, although the magnitude of the gradient changes tended to be higher among women (**Figures S60-S63** in the appendix). Furthermore, our results are similar when we consider the prevalence of elevated blood glucose and blood pressure alone (**Figures S42, S43, S49, S50** in the appendix).

### Changing urban-rural gradients

Between the NFHS 4 and 5, rural areas were adversely affected by trends in CVD risk factors compared with urban areas. In 2015-16, the prevalences of overweight, diabetes, and hypertension were 14·3, 1·4, and 3·9 percentage points higher among urban residents, respectively (**Figures S36, S40, and S57** in the appendix). However, rural areas experienced a faster increase in the prevalence of these risk factors between 2015-16 and 2019-21 (**Figures 1 and 2**): overweight increased by 4·4 percentage points (95% CI 4·1, 4·8; relative change 29·6%) in rural compared to 2·5 percentage points (95% CI 1·6, 3·5; relative change 8·6%) in urban areas; diabetes increased by 0·4 percentage points (95% CI 0·3, 0·6; relative change 15·9%) in rural compared to 0·2 percentage points (95% CI -0·2, 0·6; relative change 4·8%) in urban areas; hypertension increased by 0·7 percentage points (95% CI 0·3, 1·0; relative change 4·0%) in rural compared to a 1·6 percentage points decline (95% CI -2·3, -0·8; relative change -7·7%) in urban areas. Tobacco use was more common in rural areas in 2015-16 (urban: 22·9%; rural: 28·5%) but decreased more rapidly in urban (−5·8 percentage points, 95% CI -6·9, -4·7) compared to rural areas (−5·4 percentage points, 95% CI -6·1, -4·7) (**Figure S32** in the appendix, **Figure 1**, and **Figure 2**).

### Changing regional gradients

The prevalence of unhealthy weight, diabetes, and hypertension was highest in south India in both 2015-16 and 2019-21 (**Figure S54** in the appendix). However, regions with a relatively lower prevalence in 2015-16 had a faster increase in diabetes and hypertension between waves (**Figure 3 and Figure S55** in the appendix). For instance, central (13·7%, 95% CI 13·3, 14·0) and eastern (15·2%, 95% 14·7, 15·8) regions initially had the lowest prevalence of hypertension but were the only regions that experienced increasing prevalence over time (central 3·9 percentage points, 95% CI 3·5; 4·4; eastern: 1·2 percentage points, 95% CI 0·5, 1·8). As a result, diabetes and hypertension became more equal across regions while obesity became even higher in the south compared to the other regions.

**Figure 3.**
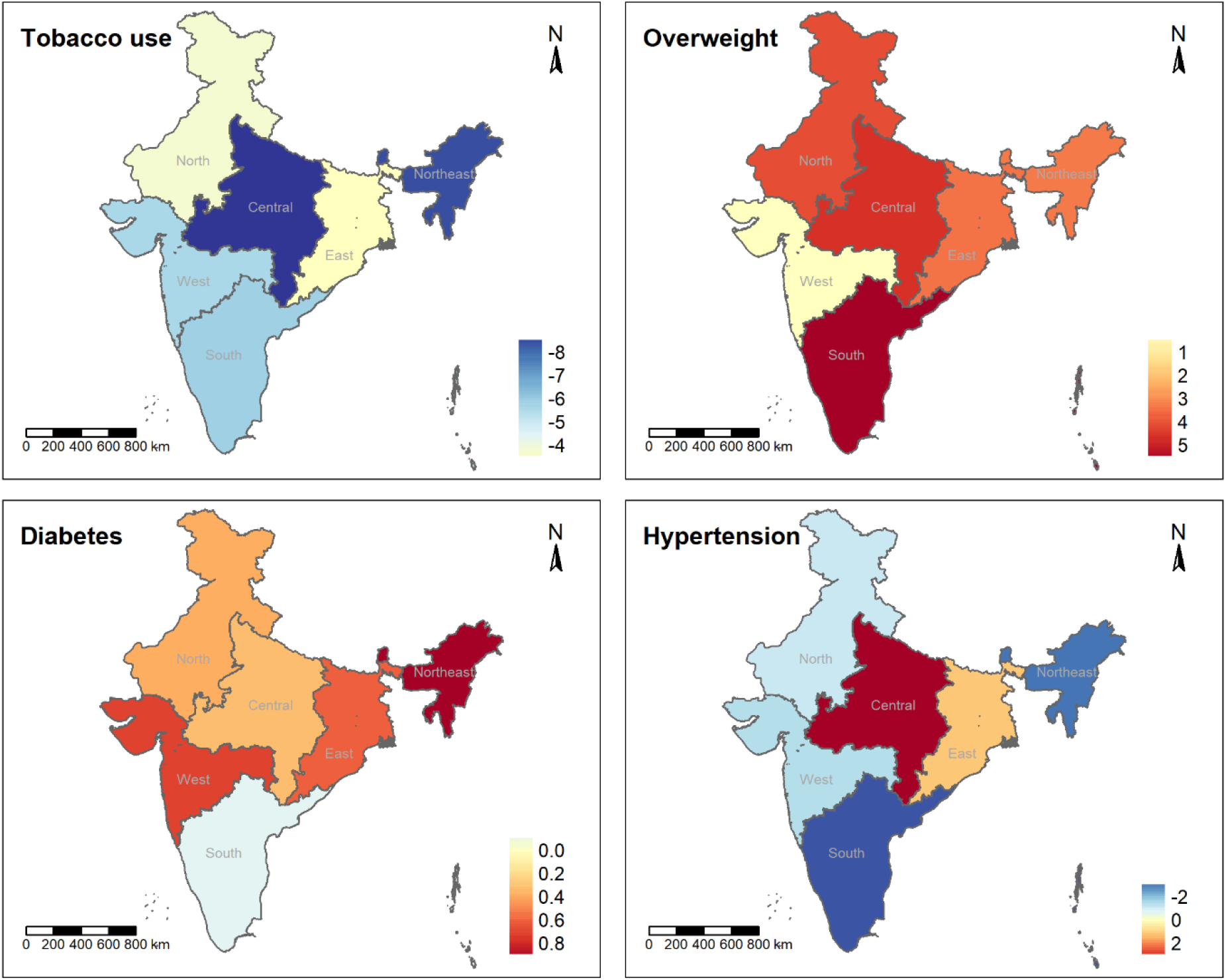
**Age-adjusted absolute changes (percentage points) in the prevalence of CVD risk factors across Indian regions, National Family Health Survey 4 (2015-16) and 5 (2019-21), India**

In 2015-16, tobacco use was lowest in southern India, with a prevalence of 17·1% (95% CI 16·3, 18·0). In contrast, central (32·6%, 95% CI 31·9, 33·3) and northeastern regions (45·3%, 95% CI 44·0, 46·7) had the highest prevalence (**Figure S54** in the appendix). These same two regions experienced the strongest absolute decline (central: -9·0 percentage points, 95% CI -10·0, -8·0; northeastern: -8·6 percentage points, 95% CI -10·4, -6·7) (**Figure 3 and Figure S55** in the appendix) while the most pronounced relative decline occurred in the south (−34·0%, 95% CI -38·6, -29·0) (**Figure S56** in the appendix). Despite heterogeneous trends across regions, as of 2019-21, the north continues to have the highest tobacco consumption and the south the lowest.

Except for tobacco consumption, adults living in EAG member states had substantially lower CVD risk factor levels in 2015-16 (**Figure S57** in the appendix). For example, overweight was more than 10 percentage points lower in the EAG (EAG: 13·8, 95% CI 13·5, 14·0; non-EAG: 24·5%, 95% CI 24·0, 25·0). Absolute and relative increases in overweight between 2015-16 and 2019-21 were higher in EAG states while these disadvantaged states were relatively less affected by increasing diabetes prevalence (EAG: 0·2 percentage points, 95%CI 0·1, 0·4; non-EAG: 0·5 percentage points, 95% CI 0·2, 0·7) (**Figures S58 and S59** in the appendix). For hypertension, the prevalence rose in EAG states (2·6 percentage points, 95%CI 2·2, 2·9) but declined in non-EAG states (−2·1 percentage points, 95% CI -2·6, -1·6). Regardless of EAG membership, tobacco use decreased over time but the decline was more pronounced in the EAG region with the initially higher prevalence (EAG: -6·7 percentage points, 95% CI -7·5, -6·0; non-EAG: -4·7 percentage points, 95%CI -5·6, -3·8). As a result of these changes, the difference in diabetes between EAG and non-EAG states slightly increased, whereas the remaining risk factors (tobacco, overweight, and hypertension) became more similar across the two groups.

## Discussion

We found that people with lower socioeconomic status (SES) measured by education and wealth and those living in rural areas experienced stronger adverse trends in unhealthy weight, hypertension, and diabetes between 2015-16 and 2019-21. While these risk factors were initially higher among higher SES and urban populations, these trends over time resulted in a substantial flatting or even reversal of SES gradients to the disadvantage of relatively poor and less educated subpopulations. For instance, the prevalence of overweight surged from around 5% to 10% among adults in the poorest wealth quintile (a relative increase of nearly 80%) compared to a change of less than 3 percentage points (relative change 8%) among adults in the richest wealth quintile. Importantly, these findings are not just the result of changes in risk factor diagnosis but rather due to secular changes in risk. The patterns we observe for unhealthy weight are based on measured height and weight and although our definitions of diabetes and hypertension consider diagnosis, we find similar trends when just focusing on measured blood glucose and blood pressure. Our results reveal that CVDs can no longer be considered a problem of the affluent parts of society and suggest that CVD prevention efforts that reach less advantaged subpopulations are needed. This need is especially pressing as lower SES groups form the majority of India’s massive population yet have much higher difficulty accessing care and treatment. ^35–37^

Our results are consistent with prior subnational studies from India, which identified early signs of reversing socioeconomic gradients in more developed settings of the country. Both urban subpopulations and more affluent states showed a higher prevalence of CVD risk factors among people with lower SES while opposite gradients prevailed elsewhere. ^5,6,18,38,39^ Our study combined with the existing literature demonstrate that changes in the social patterning of CVD risk are taking place throughout the country and at the national level.

The changing socioeconomic and geographic gradients we observe are consistent with the “reversal hypothesis” that as countries develop, CVD risk goes from being disproportionate among the rich to disproportionate among the poor. ^10,14,40^ To what extent gradients have already reversed, however, differed depending on the applied definition of SES. Measuring SES by the level of education, we find that gradients have already reversed to a large extent: unhealthy weight is the only CVD risk factor that remained higher among more educated individuals in 2019-21, while the other risk factors showed higher levels among those in the lowest education groups. In contrast, when using wealth as an indicator of SES, gradient reversal was still in the early stages: as of 2019-21, the prevalence of diabetes, hypertension, and unhealthy weight was still higher in the more affluent parts of the population, although these groups became more equal in terms of prevalence over time. These findings are in line with systematic reviews reporting a sensitivity of results to the SES definition, including the level of development at which reversal occurs. ^11,41^

As a response to alarming predictions regarding the expected future CVD burden and a corresponding threat to sustainable development, the Indian government in 2012 adopted a national action plan to prevent and control non-communicable diseases (NCD). This plan defined 9 intermediate targets for 2020 and 10 long-term targets for 2025, including a reduction of hypertension (2020: 10%, 2025: 25%), tobacco use (2020: 15%, 2025: 30%), and premature mortality from NCDs (2020: 10%, 2025: 25%) and no further increase in diabetes or obesity. ^42^ Based on our findings India not only met but exceeded the mid-term target to reduce tobacco use by 15% until 2020. The sustained decline since 2009^43^ might indicate the potential effectiveness of strong preventive efforts and serve as a promising example for other CVD risk factors: over the last decades, India introduced several laws and prevention programs to curb the disease burden caused by tobacco consumption (e.g., Cigarettes and Other Tobacco Products Act from 2003 and the National Tobacco Control Programme). ^23^ In spite of these promising trends in tobacco consumption, our results also show that changes at the national level can mask substantial adverse trends in certain subpopulations. For instance, the absence of a national change in hypertension hides the pronounced rise among low SES people. This calls for policy targets which pay attention to potentially highly vulnerable groups that make up a considerable part of the large Indian population.

Our study has some important limitations. First, only adults aged 15-49 years were eligible for the assessment of CVD risk factors in both survey rounds. As CVDs and most of their metabolic risk factors are positively correlated with age, our estimates are not nationally representative for adults aged 18 years and over and most likely underestimate the true burden, at least in terms of the year-specific levels, and are best interpreted as being relevant for younger to middle-aged adults. However, since these people will age over time, similar gradient changes in the elderly can be expected. Secondly, antidiabetic medication was not part of the NFHS-4 survey instruments so that we could not classify people based on their treatment status. While this might result in a lower diabetes prevalence, we think the difference would be marginal (most people who receive treatment will also report to have been diagnosed with diabetes and the share that actually receives treatment is low in India^44^) and that a substantial effect on trends over time is unlikely. Third, the NFHSs only measured random blood glucose and on a single occasion. While this is not the most precise diagnostic test for diabetes, ^45^ misclassification is unlikely to differ across survey rounds or to be substantially correlated with SES. South Asians tend to develop CVDs at lower values of established risk factors compared with other ethnicities, particularly BMI. ^46–50^ While some European studies with Asian expatriates determined Asian-specific BMI values that are risk-equivalent to the conventional obesity cutoff used for Caucasian populations, these thresholds might not be valid for all CVDs and might not equally apply in the Indian context. ^46^ In the absence of uncontroversial thresholds, we used the traditional overweight and obesity cutoffs. However, the traditional overweight cutoff of 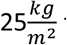 that we used for our overweight or obesity indicator is close to most of the proposed Asian-specific obesity cutoffs so that it should provide a realistic impression of people at risk. ^46,47^ Finally, the estimated changes in diabetes and hypertension may partially be driven by differential self-reporting of disease status or differential access to screenings across survey waves. However, our sensitivity analysis with alternative diabetes and hypertension indicators solely based on biomarker measurements shows that our main conclusions regarding gradient changes are robust and that secular disease trends are underlying the shift of the disease burden.

Using nationally representative data from two survey rounds with reliable biomarker measurements, our analysis provides a comprehensive picture of considerable changes in the social patterning of all major CVD risk factors. Due to disproportionate absolute and relative increases in the prevalence of unhealthy weight, hypertension, and diabetes among Indians with low SES and in rural areas since 2015-16, the socioeconomic and geographic gradients in the country substantially changed to the detriment of less-educated, relatively poor, and rural subpopulations and CVD risk factors are now far more widespread throughout the population. To prevent a severe double burden of infectious diseases and NCDs among vulnerable subpopulations and to achieve a sustainable reduction in CVD mortality, CVD prevention programs that ensure to reach marginalized population groups will be essential.

## Supporting information

Supplemental tables snd figures

STROBE checklist

## Data Availability

The data used for the analysis is publicly accessible (https://dhsprogram.com/).

## Ethics approval

This analysis of de-identified, publicly available data has received a determination of not-human subjects research by the institutional review board of the Universitätsklinikum Heidelberg. It complies with the Declaration of Helsinki.

## Consent to participate

To the best of our knowledge, the organizers of the Indian National Family and Health Surveys (NFHS-4 and NFHS-5) obtained informed consent from all survey respondents.

## Data and computing code availability

The data used for the analysis is publicly accessible (https://dhsprogram.com/). We will post our analysis code in a public repository upon publication of the manuscript.

## Author contributions

PG conceived of the study idea. SW, NS, and PS developed the methodological approach. Supervised by NS and PG, SW conducted the data analysis and wrote the initial manuscript. Both SW and NS directly accessed and verified the reported data. PG and NS acquired the necessary funding. All authors discussed the interpretation of results, contributed to revising the manuscript, and accept responsibility to submit for publication.

