## Supplemental tables snd figures for "Changing socioeconomic and geographic gradients in cardiovascular disease risk factors in India – Evidence from nationally representative household surveys"

* Joint senior authors

Corresponding author:

| | |
|---|---|
| Name: | Sarah Wetzel, MA |
| Address: | Heidelberg Institute of Global Health, Heidelberg University<br>Im Neuenheimer Feld 130.3<br>69120 Heidelberg, Germany |
| Email: | |
| Tel.: | +49 6221 56-5344 |
| Fax.: | +49 6221 56-5948 |
| ORCID: | 0000-0002-7376-1770 |

### Contents

Sampling procedure ... 3
Definitions ... 4
Age-adjusted and weighted sample characteristics ... 5
Trends at the national level ... 6
Estimates using the time-specific age-distribution ... 6
Age-adjusted estimates ... 6
Results stratified by socioeconomic group or place of residence ... 7
Estimates using the time-specific age-distribution ... 7
Tobacco consumption ... 7
Cigarette smoking ... 9
Overweight or obesity ... 10
Obesity ... 11
Diabetes ... 12
High blood glucose ... 13
Self-reported diabetes ... 14
Hypertension ... 15
High blood pressure ... 16
Told to have high blood pressure ... 17
Age-adjusted ... 18
Tobacco consumption ... 18
Cigarette smoking ... 19
Overweight or obesity ... 20
Obesity ... 21
Diabetes ... 22
High blood glucose ... 23
Self-reported diabetes ... 24
Hypertension ... 25
High blood pressure ... 26
Told to have high blood pressure ... 27
Results stratified by region ... 28
NFHS-4 ... 28
Absolute changes ... 29
Relative changes ... 30
Results stratified by regional level of development ... 31
NFHS-4 ... 31
Absolute changes ... 32
Relative changes ... 33
Results stratified by sex and socioeconomic status or place of residence ... 34
Women ... 34
Men ... 35
References ... 36

#### Sampling procedure

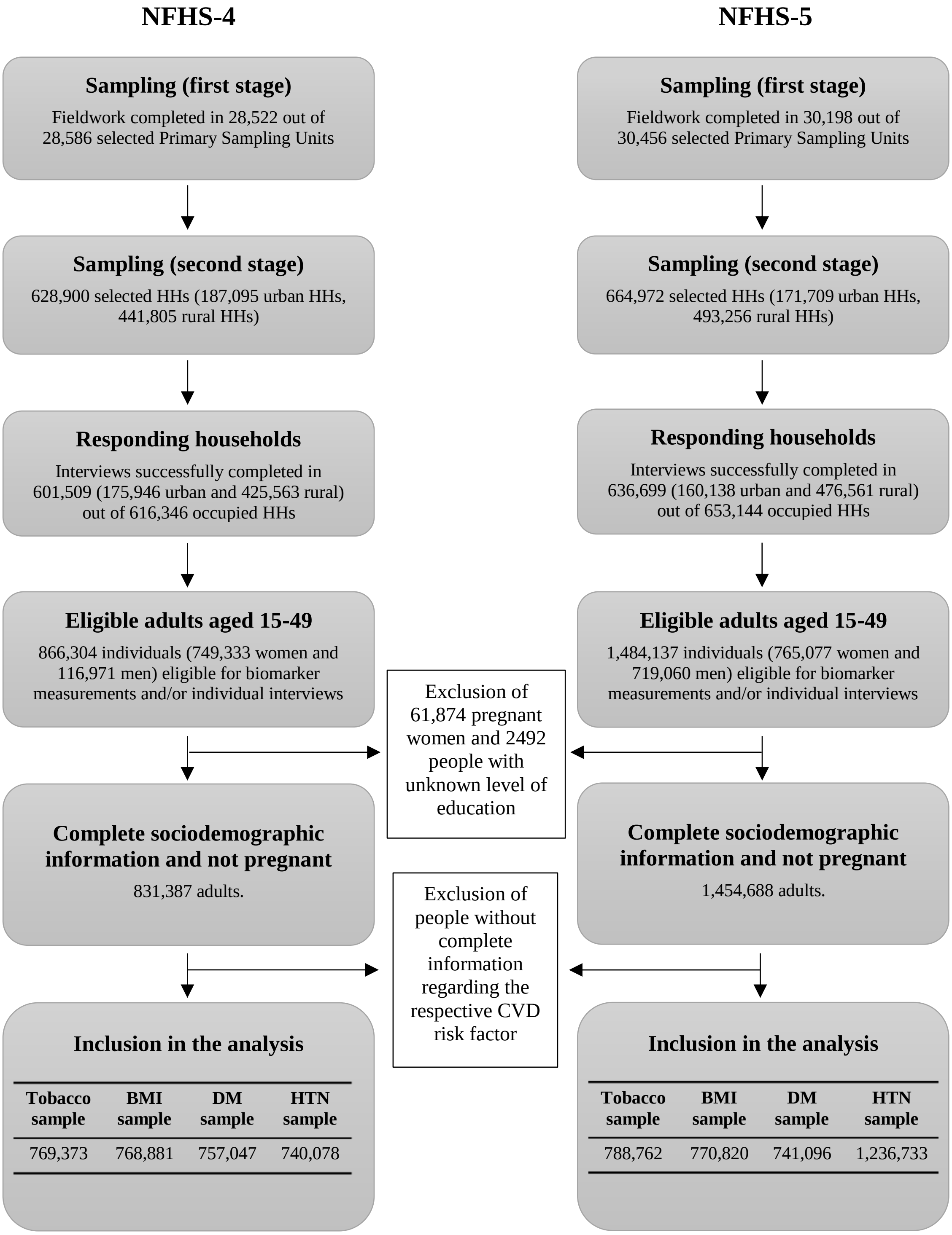


**Figure S1. Flowchart of the sampling procedure**
HH=household, BMI=body mass index, DM=diabetes, HTN=hypertension. In the NFHS-5, adults aged 15 years and older were eligible for glucose tests as well as for blood pressure measurements and asked about hypertension diagnoses or treatments while all other mentioned CVD-related items only covered men aged 15-54 in a representative subsample of 15% of households (state module) and women aged 15-49 years.

##### Definitions

As mentioned in the discussion, the share of the population that had been diagnosed with hypertension followed a strong and potentially unrealistic downward trend in a few states which might indicate potential data quality issues with regard to this variable. To demonstrate the robustness of our findings, particularly the reversal of socioeconomic gradients, we present results for three different hypertension indicators below: i) an indicator for an average systolic blood pressure greater than or equal to 140mmHg or an average diastolic blood pressure greater than or equal to 90mmHg, ii) an indicator for being told to have high blood pressure by a clinician on two different occasions, and iii) the combined indicator defined as previously (high blood pressure, told to have high blood pressure, or current medication). Similarly, we show trends for three different diabetes indicators to show the contribution of secular disease trends, trends in self-reporting due to changing screening behavior or access to care, and the combined effect of the two: i) an indicator for a random plasma glucose concentration $\geq 200\frac{mg}{dL}$, ii) an indicator for self-reported diabetes status, and iii) the combined indicator defined as previously (high blood glucose or self-reported diabetes). We measured cigarette smoking as a binary indicator based on self-report. All other variables are defined as before.

To assess the contribution of aging, we compare age-adjusted estimates (direct age-standardization based on the WHO standard population [1]) with the estimates using the actual time-specific age distribution of the Indian population. To ensure that our data was representative of the age distribution of the Indian population aged 15-49 after applying the exclusion criteria, we adapted the household weights based on the Indian age distribution in the respective survey year according to the United Nations World Population Prospects population estimates (UNWPP). [2] However, mean age among adults aged 15-49 increased by less than 0.4 years between survey rounds. As a result, age standardization based on the WHO standard population only had a minor impact on our estimates and the conclusions regarding the trends in CVD gradients hold true for both crude and age-adjusted estimated.

#### Age-adjusted and weighted sample characteristics

| | Smoking sample | | Overweight sample | | Diabetes sample | | Hypertension sample | |
|---|---|---|---|---|---|---|---|---|
| | **NFHS-4** *N = 769,373* | **NFHS-5** *N = 788,762* | **NFHS-4** *N = 768,881* | **NFHS-5** *N = 770,820* | **NFHS-4** *N = 740,078* | **NFHS-5** *N = 741,096* | **NFHS-4** *N = 757,047* | **NFHS-5** *N = 1,236,733* |
| **Sex** | | | | | | | | |
| Male | 388,960 (51%) | 369,217 (47%) | 387,513 (50%) | 357,005 (46%) | 372,337 (50%) | 341,805 (46%) | 381,198 (50%) | 572,340 (46%) |
| Female | 380,413 (49%) | 419,545 (53%) | 381,368 (50%) | 413,815 (54%) | 367,741 (50%) | 399,291 (54%) | 375,849 (50%) | 664,393 (54%) |
| **Age group** | | | | | | | | |
| 15-19 years | 125,295 (16%) | 128,452 (16%) | 125,215 (16%) | 125,531 (16%) | 120,524 (16%) | 120,690 (16%) | 123,288 (16%) | 201,406 (16%) |
| 20-24 years | 121,597 (16%) | 124,661 (16%) | 121,519 (16%) | 121,825 (16%) | 116,967 (16%) | 117,128 (16%) | 119,649 (16%) | 195,461 (16%) |
| 25-29 years | 117,307 (15%) | 120,263 (15%) | 117,232 (15%) | 117,527 (15%) | 112,840 (15%) | 112,995 (15%) | 115,427 (15%) | 188,566 (15%) |
| 30-34 years | 112,573 (15%) | 115,410 (15%) | 112,501 (15%) | 112,785 (15%) | 108,287 (15%) | 108,436 (15%) | 110,770 (15%) | 180,956 (15%) |
| 35-39 years | 105,768 (14%) | 108,434 (14%) | 105,701 (14%) | 105,967 (14%) | 101,741 (14%) | 101,881 (14%) | 104,074 (14%) | 170,018 (14%) |
| 40-44 years | 97,484 (13%) | 99,941 (13%) | 97,422 (13%) | 97,668 (13%) | 93,773 (13%) | 93,902 (13%) | 95,923 (13%) | 156,702 (13%) |
| 45-49 years | 89,348 (12%) | 91,600 (12%) | 89,291 (12%) | 89,516 (12%) | 85,946 (12%) | 86,065 (12%) | 87,917 (12%) | 143,624 (12%) |
| **Level of education** | | | | | | | | |
| No education | 149,087 (19%) | 126,440 (16%) | 149,908 (19%) | 125,129 (16%) | 144,425 (20%) | 119,960 (16%) | 148,165 (20%) | 202,255 (16%) |
| Incomplete primary | 50,361 (6.5%) | 46,325 (5.9%) | 50,598 (6.6%) | 45,813 (5.9%) | 48,482 (6.6%) | 43,838 (5.9%) | 49,754 (6.6%) | 71,727 (5.8%) |
| Complete primary | 49,271 (6.4%) | 50,177 (6.4%) | 49,643 (6.5%) | 49,376 (6.4%) | 47,674 (6.4%) | 47,457 (6.4%) | 49,182 (6.5%) | 79,365 (6.4%) |
| Incomplete secondary | 326,082 (42%) | 338,876 (43%) | 326,072 (42%) | 332,354 (43%) | 313,956 (42%) | 320,049 (43%) | 320,837 (42%) | 530,749 (43%) |
| Complete secondary | 77,454 (10%) | 88,790 (11%) | 77,521 (10%) | 86,297 (11%) | 74,418 (10%) | 83,236 (11%) | 76,306 (10%) | 139,393 (11%) |
| Higher than secondary | 117,117 (15%) | 138,155 (18%) | 115,138 (15%) | 131,851 (17%) | 111,123 (15%) | 126,556 (17%) | 112,804 (15%) | 213,244 (17%) |
| **Wealth quintile** | | | | | | | | |
| Poorest | 125,386 (16%) | 140,799 (18%) | 127,006 (17%) | 139,930 (18%) | 121,816 (16%) | 133,428 (18%) | 125,611 (17%) | 224,498 (18%) |
| Poorer | 148,512 (19%) | 158,592 (20%) | 150,247 (20%) | 157,186 (20%) | 144,451 (20%) | 150,997 (20%) | 148,450 (20%) | 248,612 (20%) |
| Middle | 161,386 (21%) | 163,912 (21%) | 161,670 (21%) | 161,722 (21%) | 155,634 (21%) | 156,126 (21%) | 159,353 (21%) | 260,865 (21%) |
| Richer | 166,149 (22%) | 165,700 (21%) | 164,990 (21%) | 161,800 (21%) | 159,172 (22%) | 155,803 (21%) | 161,975 (21%) | 261,982 (21%) |
| Richest | 167,941 (22%) | 159,759 (20%) | 164,967 (21%) | 150,182 (19%) | 159,005 (21%) | 144,742 (20%) | 161,658 (21%) | 240,776 (19%) |
| **Place of residence** | | | | | | | | |
| Urban | 273,751 (36%) | 258,811 (33%) | 268,525 (35%) | 245,630 (32%) | 258,478 (35%) | 236,048 (32%) | 263,049 (35%) | 396,768 (32%) |
| Rural | 495,622 (64%) | 529,951 (67%) | 500,356 (65%) | 525,190 (68%) | 481,600 (65%) | 505,048 (68%) | 493,998 (65%) | 839,965 (68%) |

**Table S1. Age-adjusted (WHO standard population) and weighted sample characteristics**

#### Trends at the national level

#### Estimates using the time-specific age-distribution

| Outcome | NFHS-4 | NFHS-5 | Absolute change (percentage points) | Relative change (%) |
|---|---|---|---|---|
| Tobacco Consumption | 26 [25.5; 26.4] | 20.9 [20.5; 21.3] | -5.1 [-5.7; -4.5] | -19.6 [-21.6; -17.5] |
| Cigarette smoking | 6.9 [6.7; 7.2] | 5.9 [5.7; 6.2] | -1 [-1.3; -0.6] | -14.2 [-18.8; -9.5] |
| Overweight or obesity | 19.3 [19; 19.6] | 23 [22.8; 23.3] | 3.7 [3.3; 4.1] | 19.2 [16.8; 21.6] |
| Obesity | 3.9 [3.8; 4] | 5.2 [5; 5.3] | 1.2 [1.1; 1.4] | 31.2 [26.4; 36.2] |
| Diabetes | 3.1 [3; 3.2] | 3.5 [3.4; 3.6] | 0.4 [0.2; 0.6] | 12.8 [7.6; 18.3] |
| High blood glucose | 1.9 [1.8; 2] | 2.2 [2.2; 2.3] | 0.3 [0.2; 0.4] | 16.8 [10.2; 23.6] |
| Self-reported diabetes | 1.7 [1.6; 1.8] | 1.9 [1.8; 2] | 0.2 [0.1; 0.3] | 12.9 [5.6; 20.8] |
| Hypertension | 17.1 [16.9; 17.4] | 17.3 [17.1; 17.5] | 0.2 [-0.2; 0.5] | 0.9 [-1; 2.9] |
| High blood pressure | 10.9 [10.7; 11.1] | 13.1 [13; 13.2] | 2.2 [2; 2.5] | 20.5 [18.1; 22.9] |
| Told to have high blood pressure | 7.7 [7.4; 8] | 5.4 [5.3; 5.5] | -2.3 [-2.6; -2] | -29.7 [-32.5; -26.8] |

**Table S2. Trends in CVD risk factors at the national level**

#### Age-adjusted estimates

| Outcome | NFHS-4 | NFHS-5 | Absolute change (percentage points) | Relative change (%) |
|---|---|---|---|---|
| Cigarette smoking | 7 [6.7; 7.2] | 6 [5.7; 6.2] | -1 [-1.4; -0.7] | -14.6 [-19.1; -9.8] |
| Obesity | 4.1 [4; 4.2] | 5.2 [5.1; 5.4] | 1.2 [1; 1.3] | 28.4 [23.7; 33.1] |
| High blood glucose | 2.1 [2; 2.1] | 2.3 [2.2; 2.4] | 0.3 [0.1; 0.4] | 12.9 [6.7; 19.6] |
| Self-reported diabetes | 1.8 [1.7; 1.9] | 2 [1.9; 2] | 0.2 [0; 0.3] | 9.5 [2.5; 17.1] |
| High blood pressure | 11.4 [11.2; 11.6] | 13.4 [13.2; 13.5] | 2 [1.8; 2.2] | 17.5 [15.2; 19.8] |
| Told to have high blood pressure | 8 [7.7; 8.2] | 5.5 [5.4; 5.6] | -2.4 [-2.7; -2.1] | -30.5 [-33.2; -27.6] |

**Table S3. Age-adjusted trends in CVD risk factors at the national level**

##### Results stratified by socioeconomic group or place of residence

##### Estimates using the time-specific age-distribution

##### Tobacco consumption

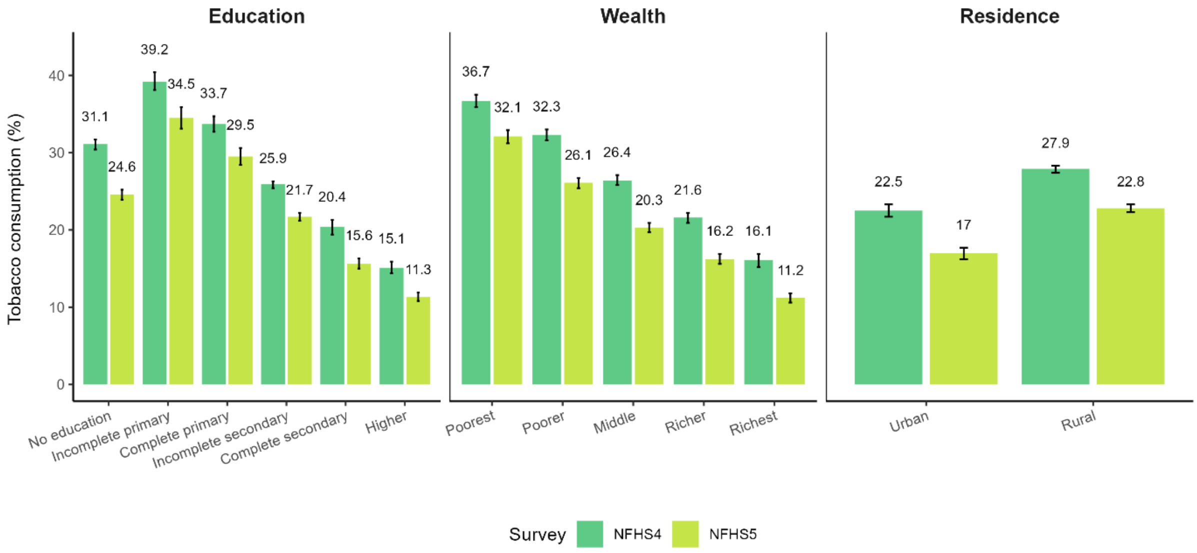


**Figure S2. Prevalence of tobacco consumption among adults aged 15-49 across subpopulations in each survey round**

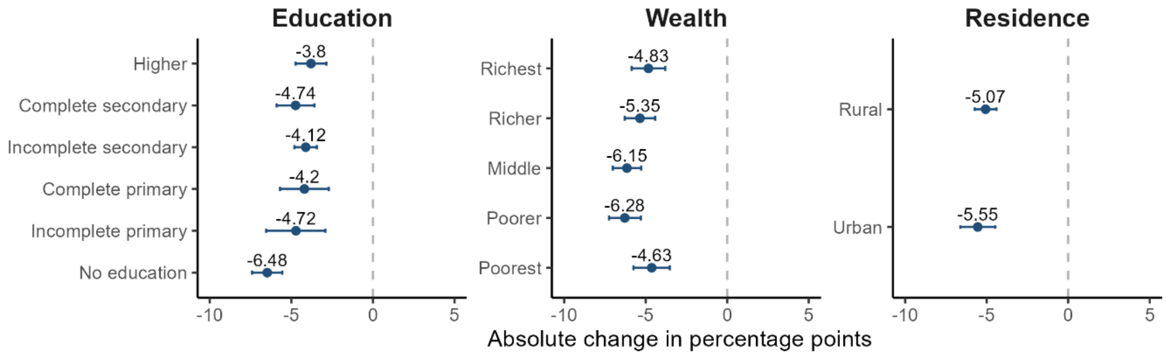


**Figure S3. Absolute changes in the prevalence of tobacco consumption among adults aged 15-49 across subpopulations**

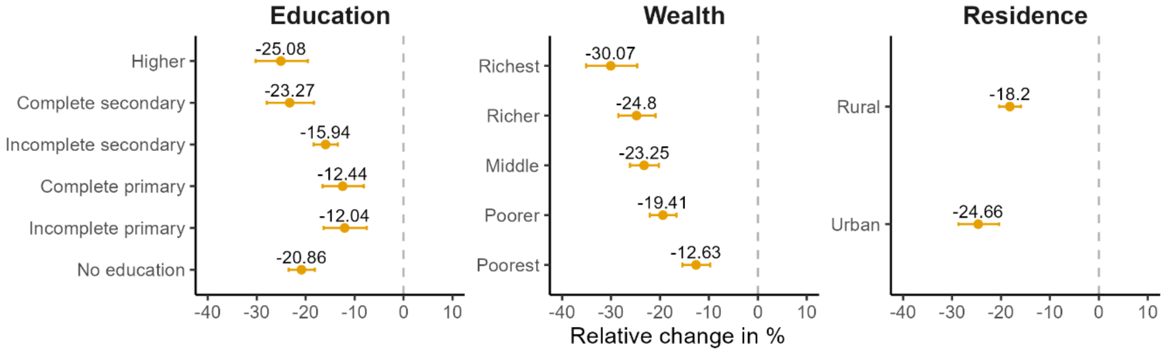


**Figure S4. Relative changes in the prevalence of tobacco consumption among adults aged 15-49 across subpopulations**

#### Cigarette smoking

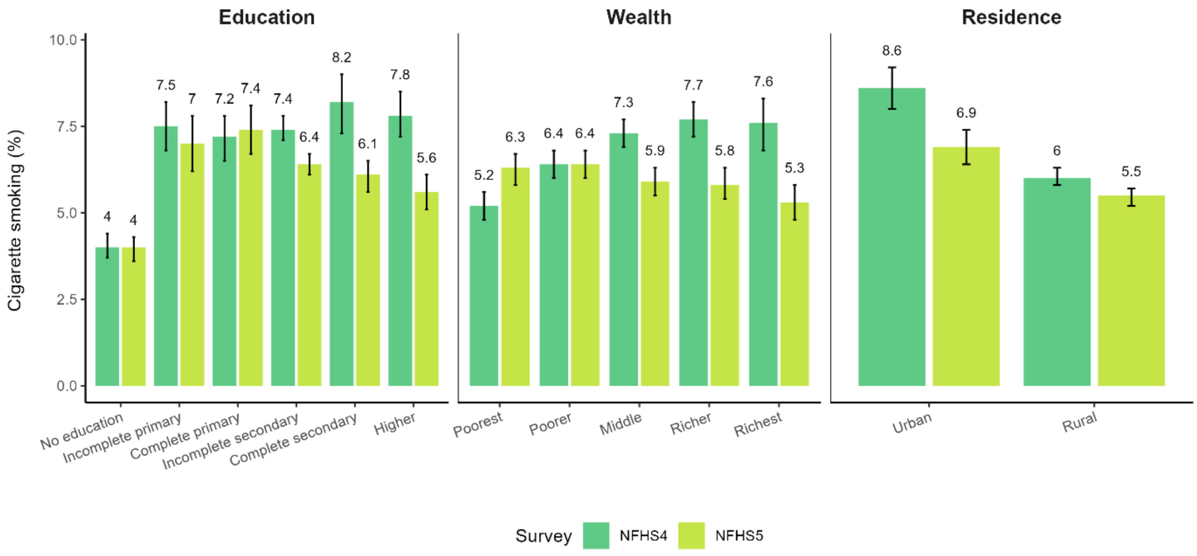


**Figure S5. Prevalence of cigarette smoking among adults aged 15-49 across subpopulations in each survey round**

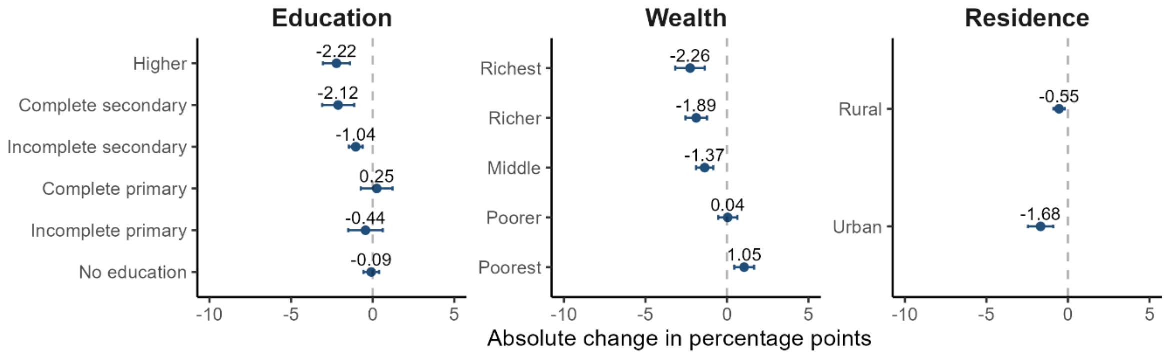


**Figure S6. Absolute changes in the prevalence of cigarette smoking among adults aged 15-49 across subpopulations**

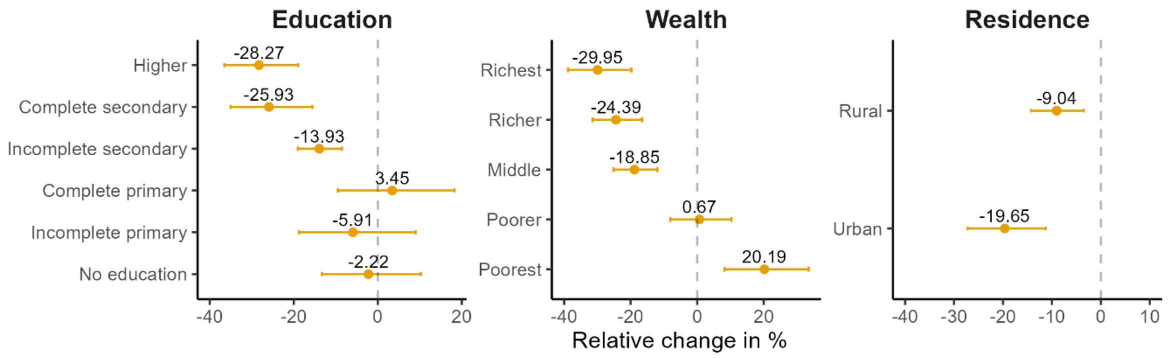


**Figure S7. Relative changes in the prevalence of cigarette smoking among adults aged 15-49 across subpopulations**

#### Overweight or obesity

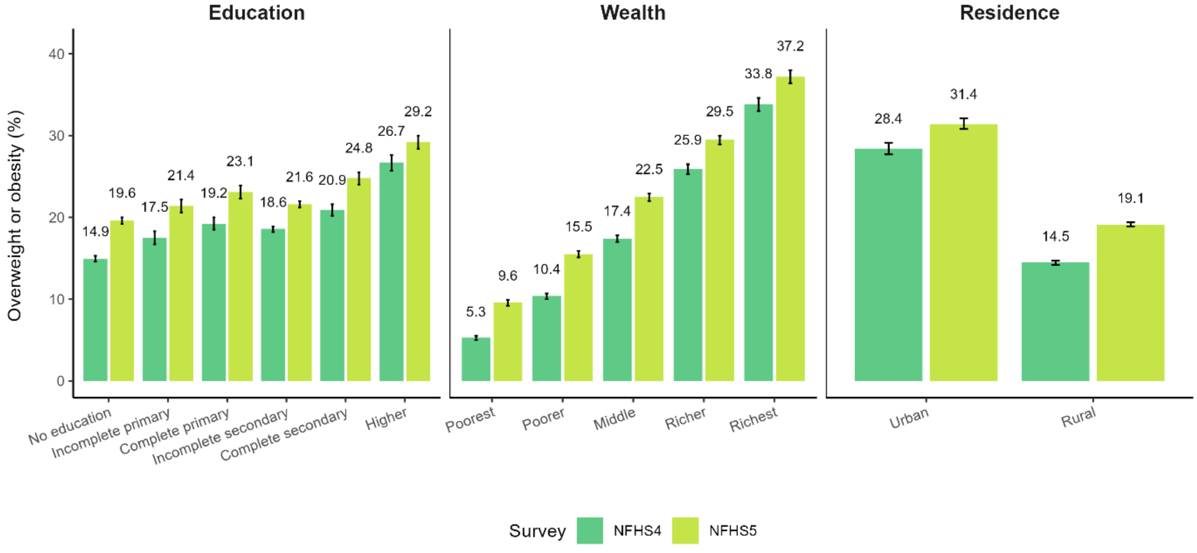


**Figure S8. Prevalence of overweight or obesity among adults aged 15-49 across subpopulations in each survey round**

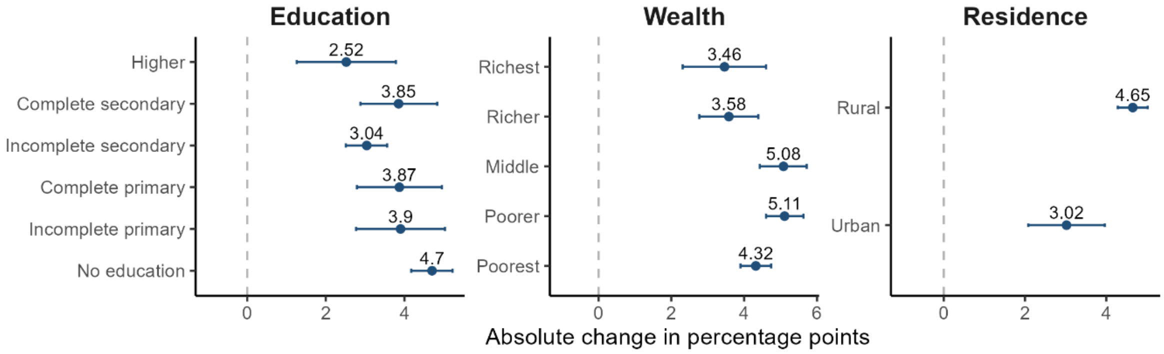


**Figure S9. Absolute changes in the prevalence of overweight or obesity among adults aged 15-49 across subpopulations**

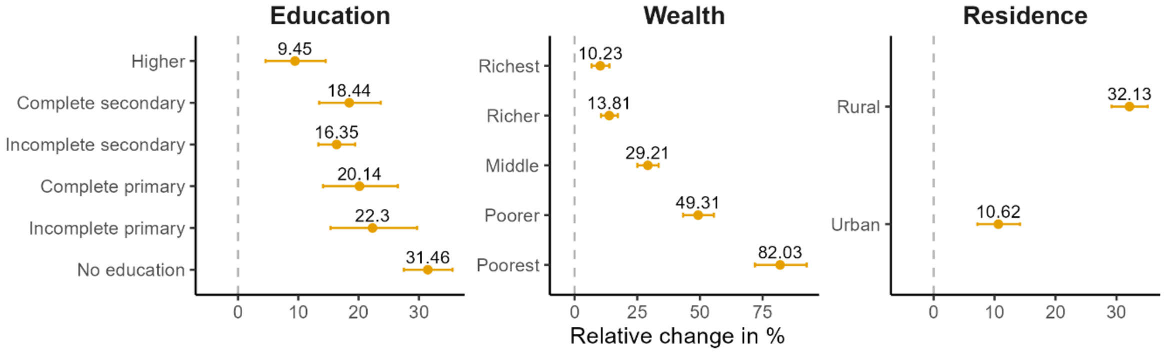


**Figure S10. Relative changes in the prevalence of overweight or obesity among adults aged 15-49 across subpopulations**

#### Obesity

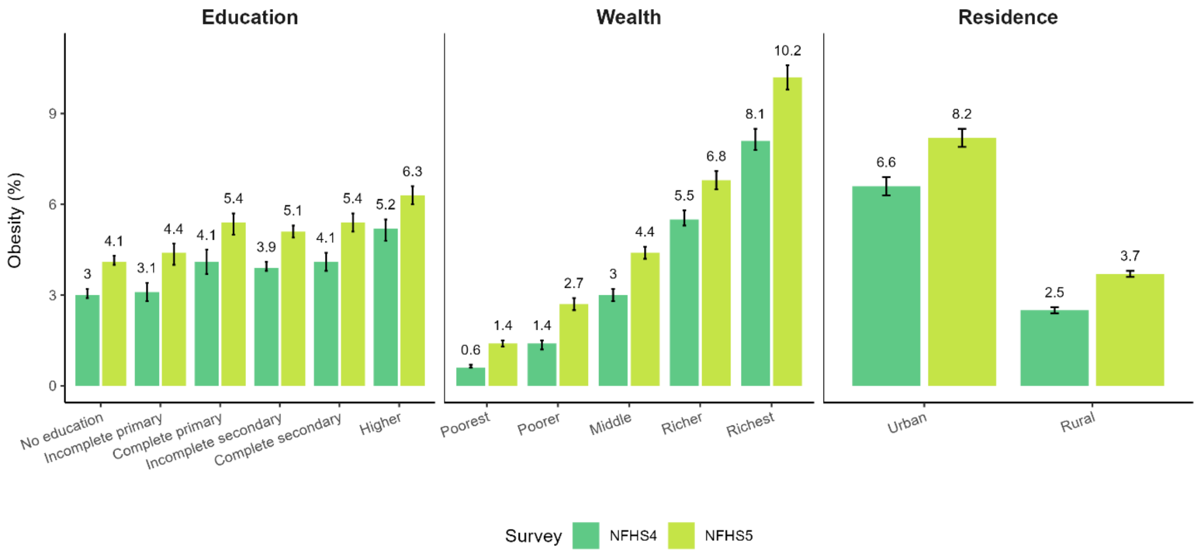


**Figure S11. Prevalence of obesity among adults aged 15-49 across subpopulations in each survey round**

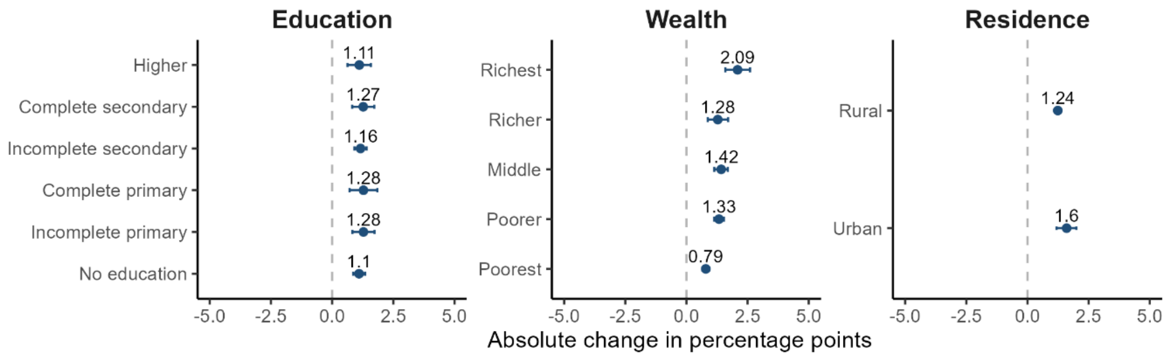


**Figure S12. Absolute changes in the prevalence of obesity among adults aged 15-49 across subpopulations**

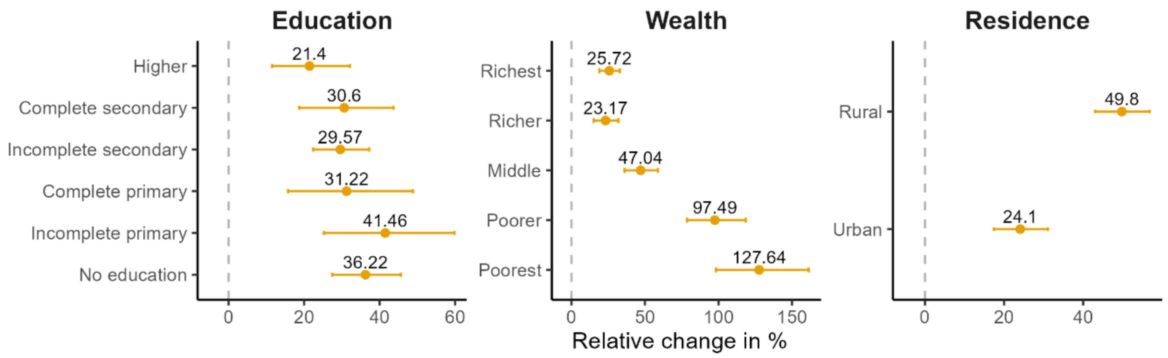


**Figure S13. Relative changes in the prevalence of obesity among adults aged 15-49 across subpopulations**

#### Diabetes

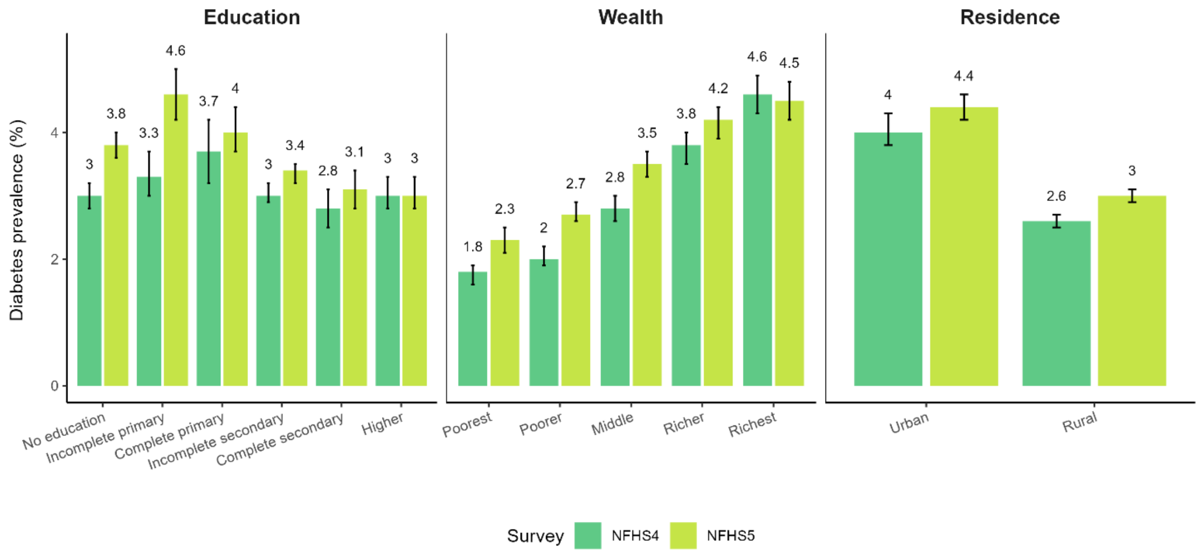


**Figure S14. Prevalence of diabetes among adults aged 15-49 across subpopulations in each survey round**

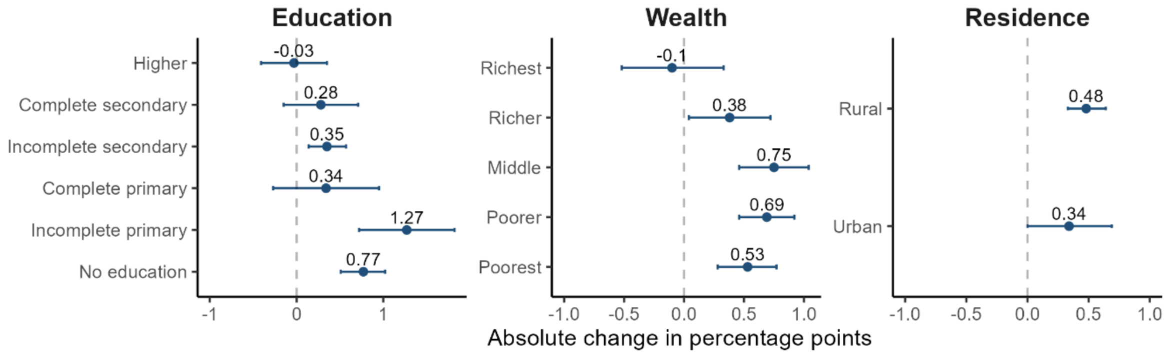


**Figure S15. Absolute changes in the prevalence of diabetes among adults aged 15-49 across subpopulations**

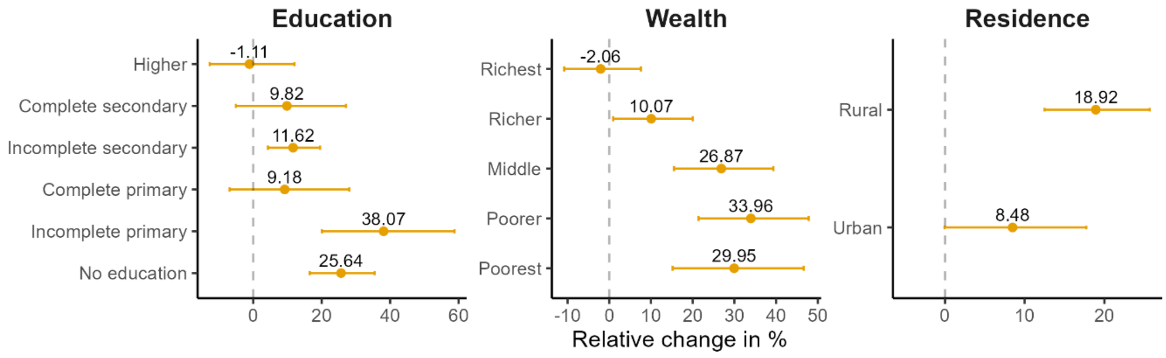


**Figure S16. Relative changes in the prevalence of diabetes among adults aged 15-49 across subpopulations**

#### High blood glucose

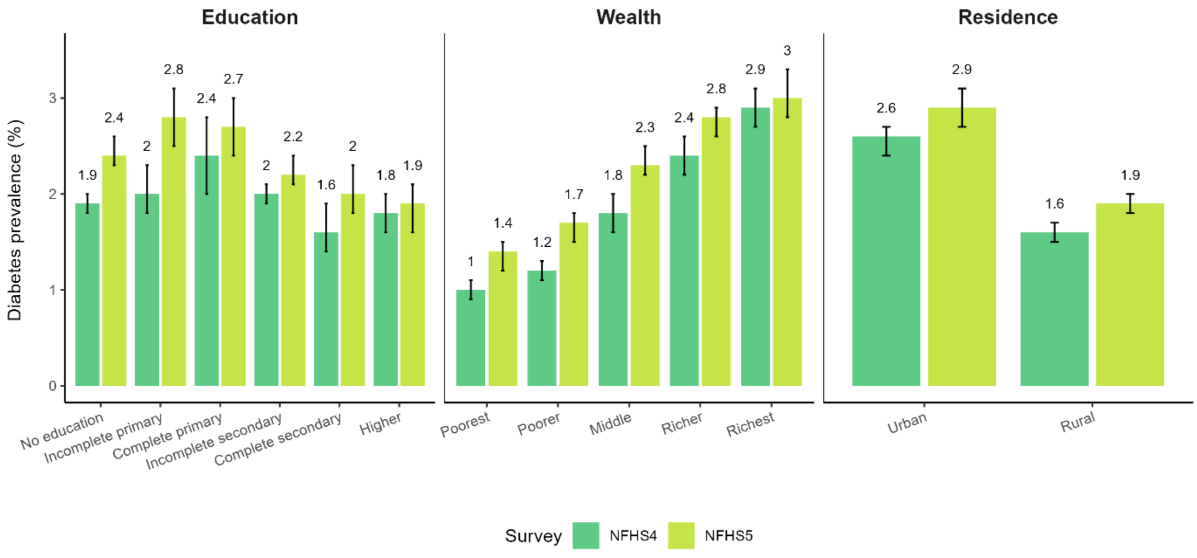


**Figure S17. Prevalence of high blood glucose among adults aged 15-49 across subpopulations in each survey round**

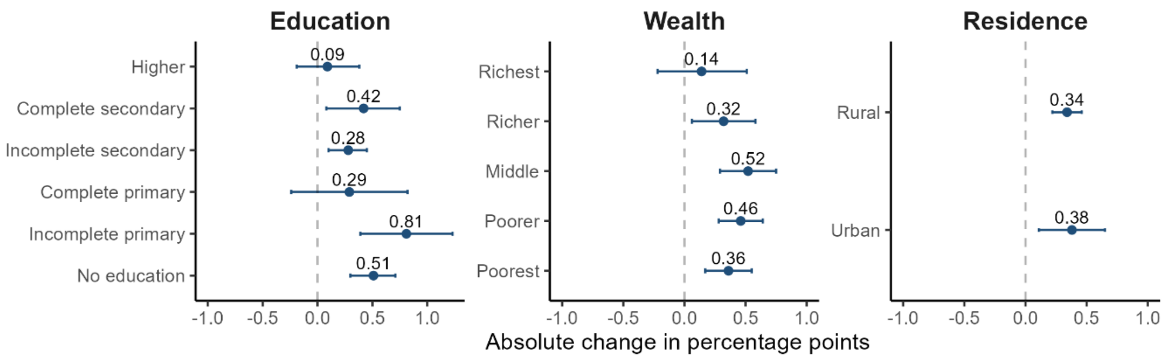


**Figure S18. Absolute changes in the prevalence of high blood glucose among adults aged 15-49 across subpopulations**

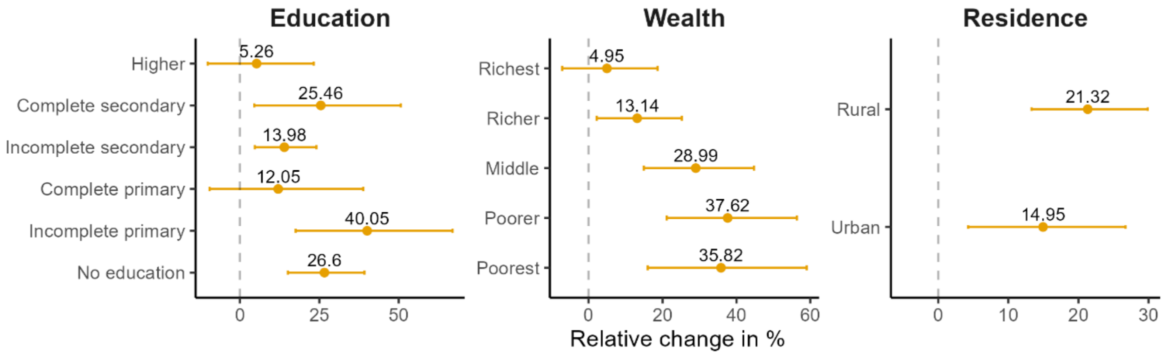


**Figure S19. Relative changes in the prevalence of high blood glucose among adults aged 15-49 across subpopulations**

#### Self-reported diabetes

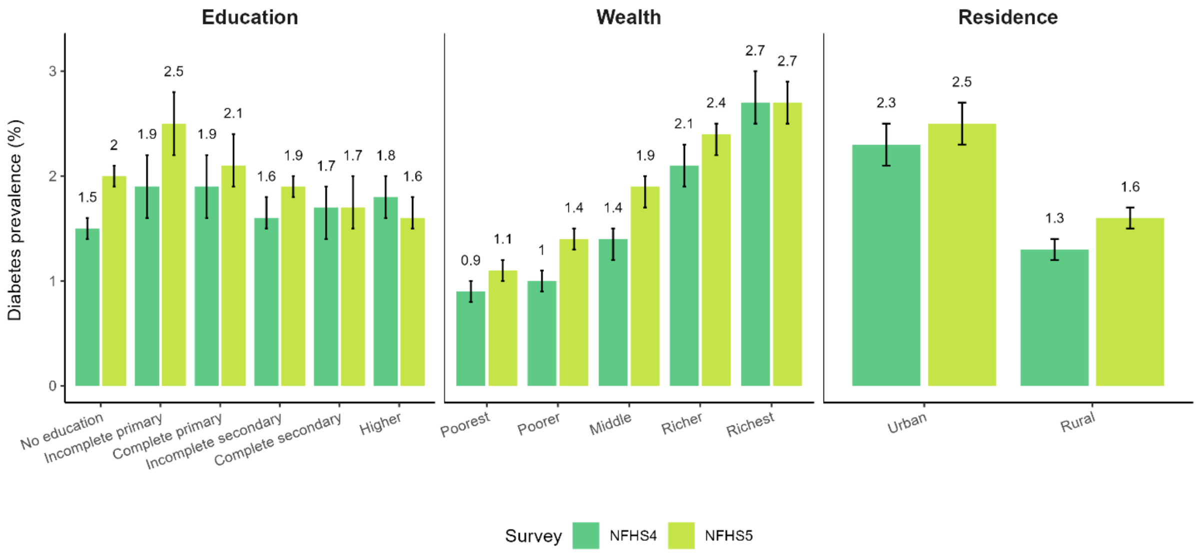


**Figure S20. Prevalence of diabetes among adults aged 15-49 across subpopulations in each survey round**

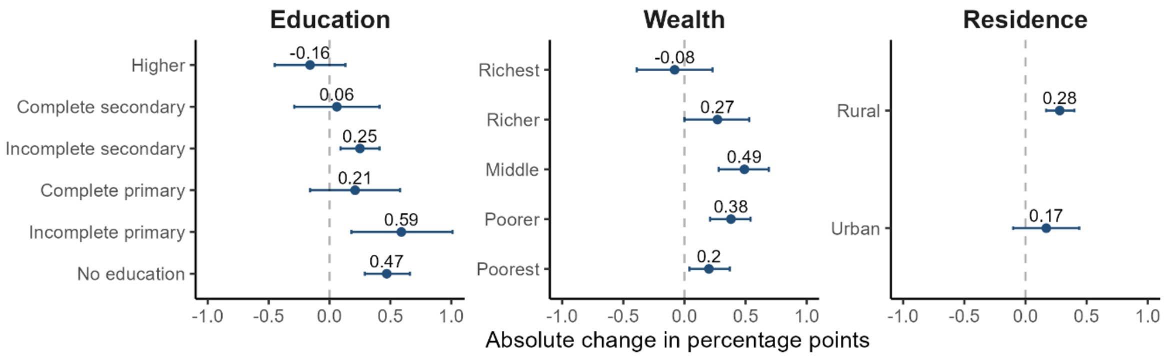


**Figure S21. Absolute changes in the prevalence of self-reported diabetes among adults aged 15-49 across subpopulations**

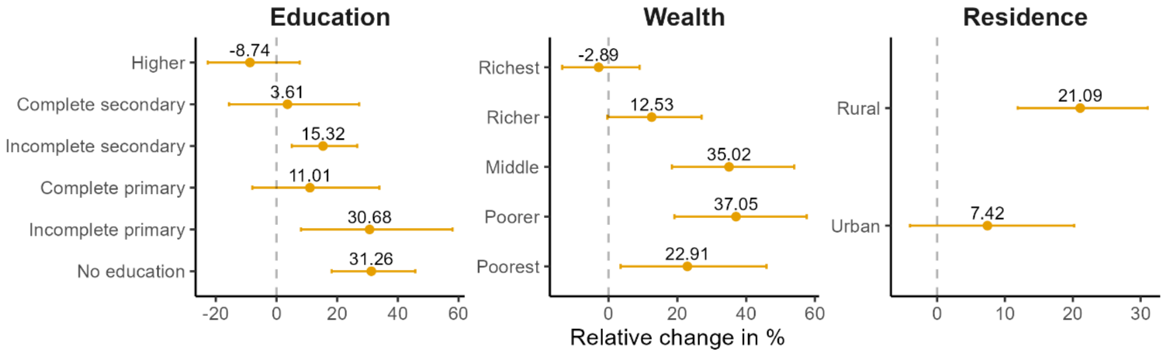


**Figure S22. Relative changes in the prevalence of self-reported diabetes among adults aged 15-49 across subpopulations**

#### Hypertension

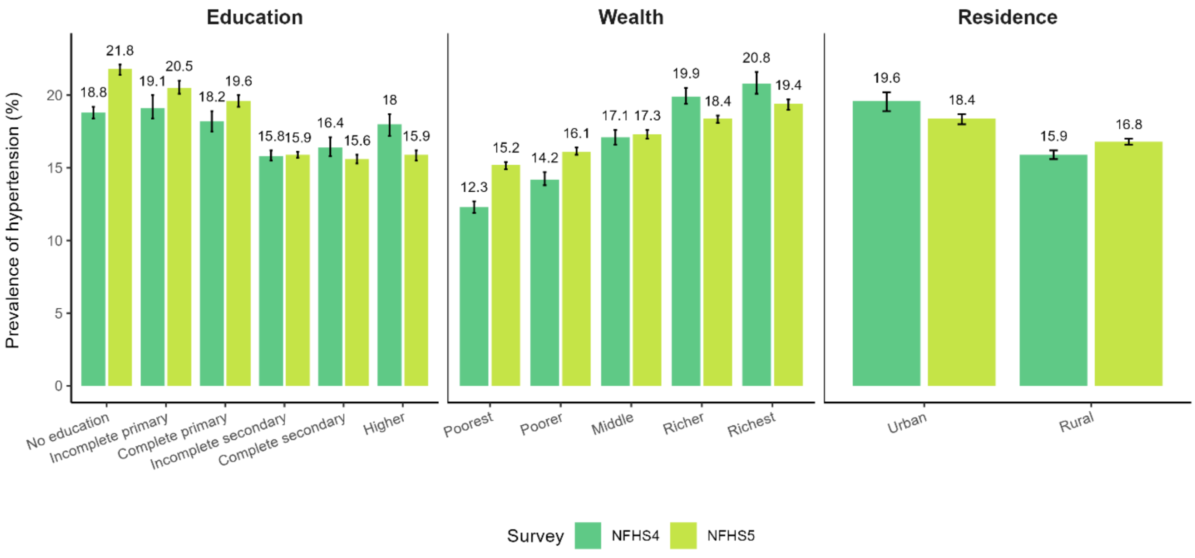


**Figure S23. Prevalence of hypertension among adults aged 15-49 across subpopulations in each survey round**

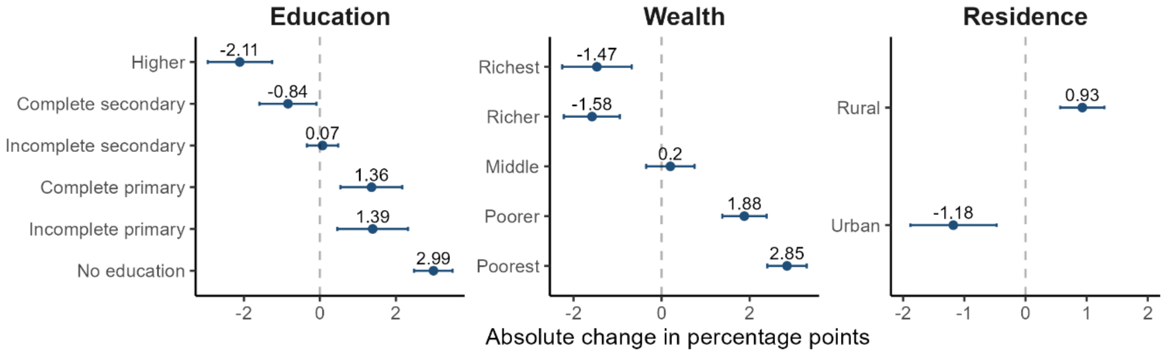


**Figure S24. Absolute changes in the prevalence of hypertension among adults aged 15-49 across subpopulations**

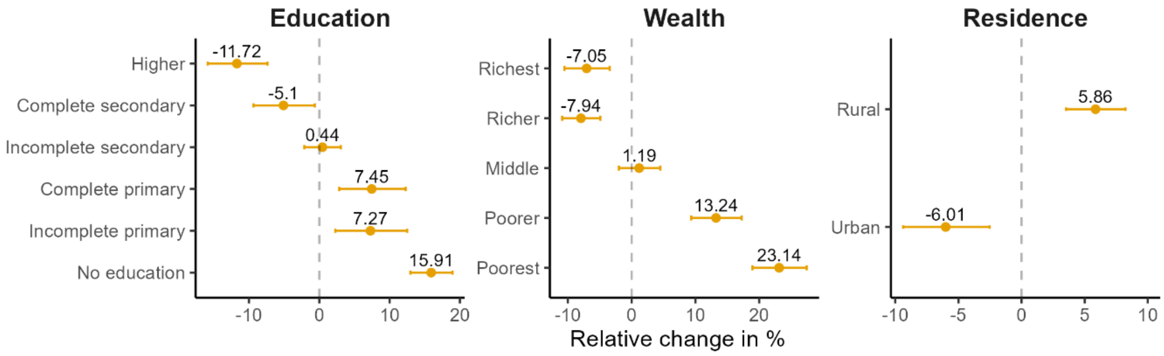


**Figure S25. Relative changes in the prevalence of hypertension among adults aged 15-49 across subpopulations**

#### High blood pressure

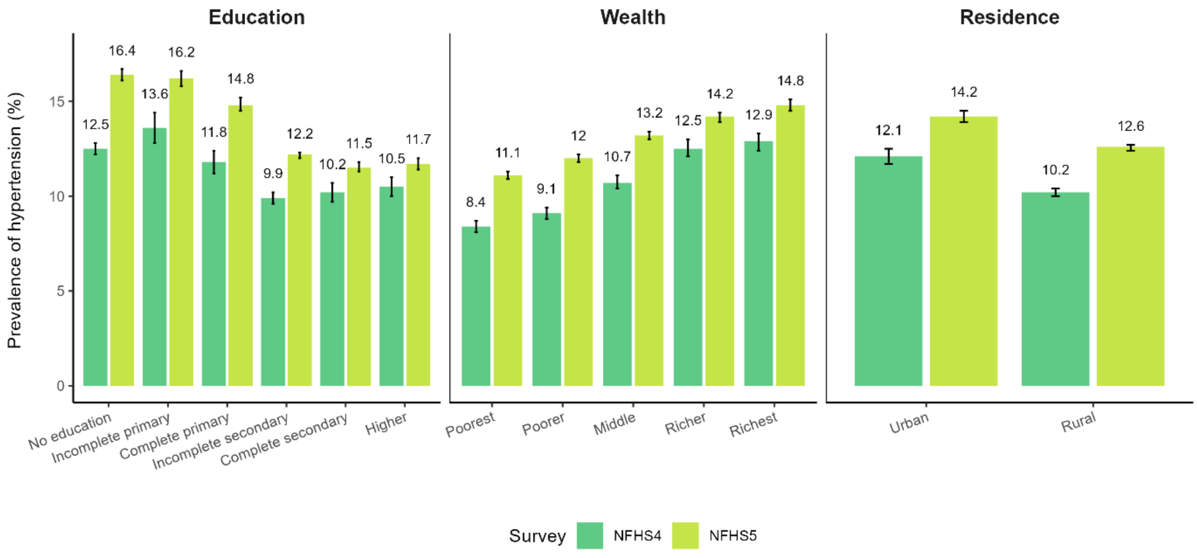


**Figure S26. Prevalence of high blood pressure among adults aged 15-49 across subpopulations in each survey round**

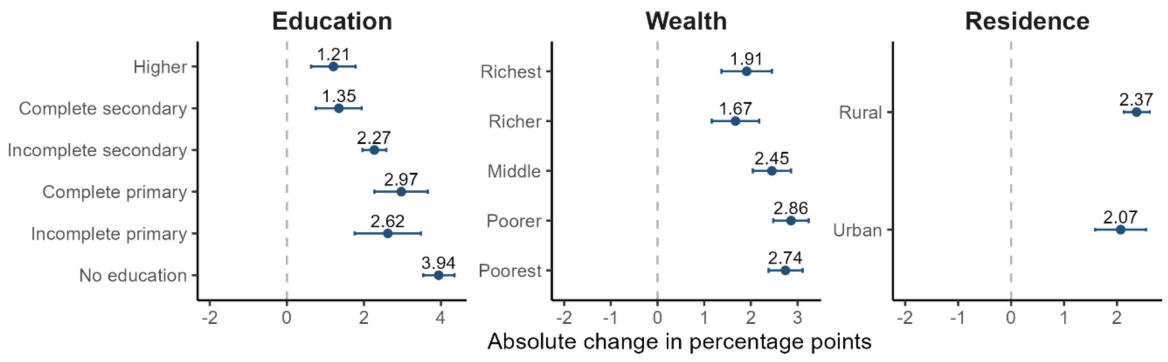


**Figure S27. Absolute changes in the prevalence of high blood pressure among adults aged 15-49 across subpopulations**

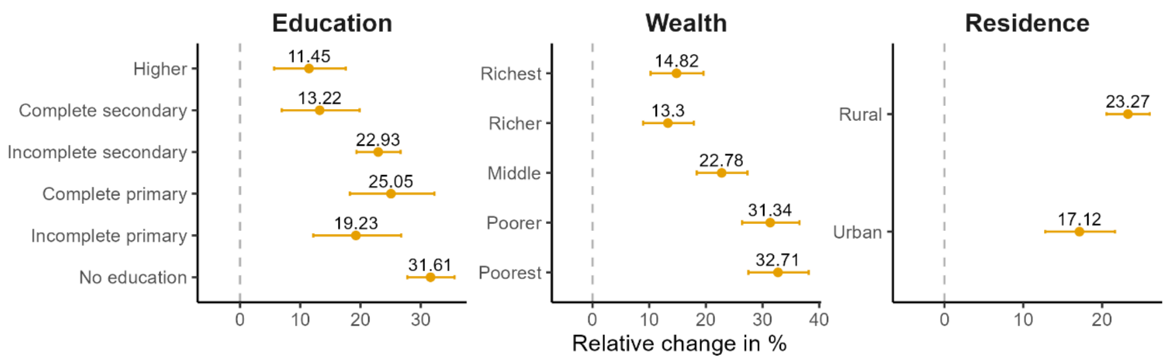


**Figure S28. Relative changes in the prevalence of high blood pressure among adults aged 15-49 across subpopulations**

#### Told to have high blood pressure

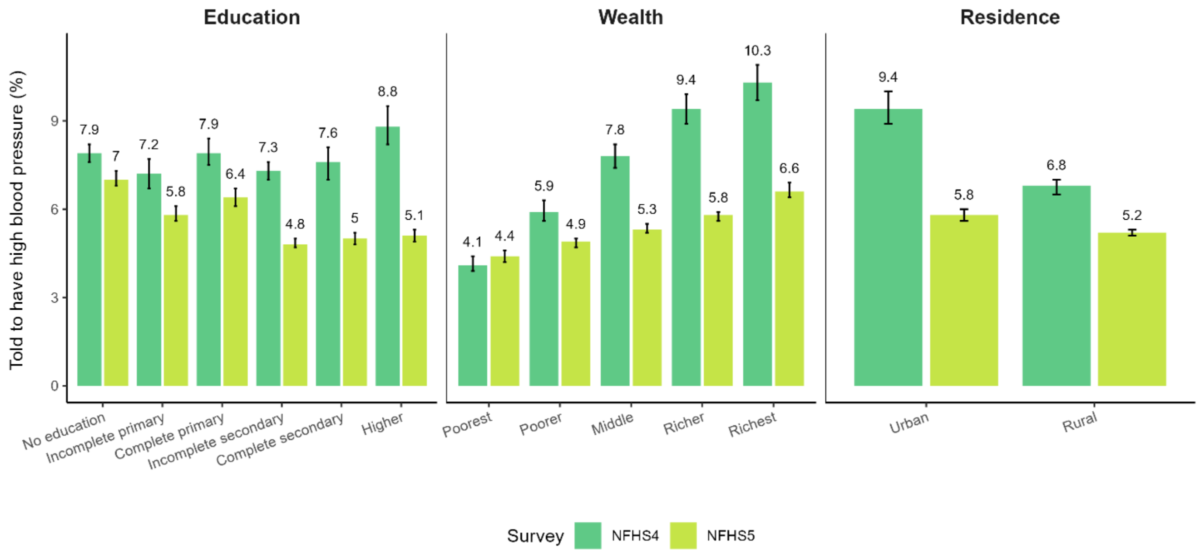


**Figure S29. Prevalence of adults aged 15-49 diagnosed with hypertension across subpopulations in each survey round**

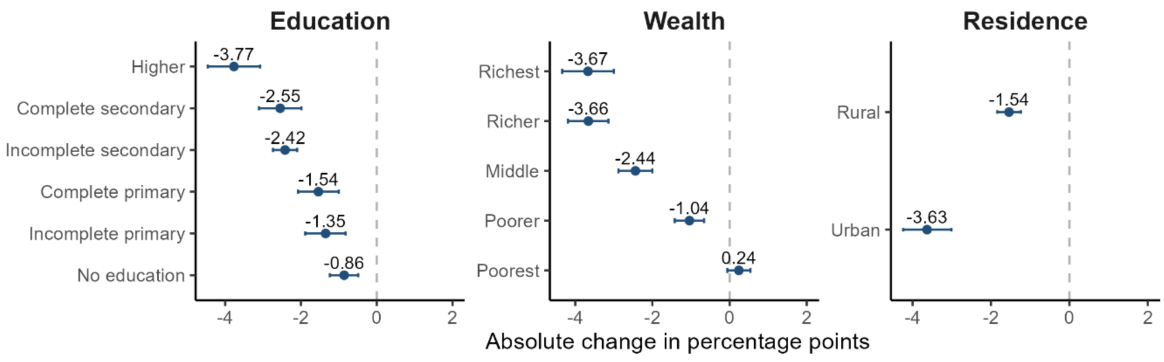


**Figure S30. Absolute changes in the prevalence of adults aged 15-49 diagnosed with hypertension across subpopulations**

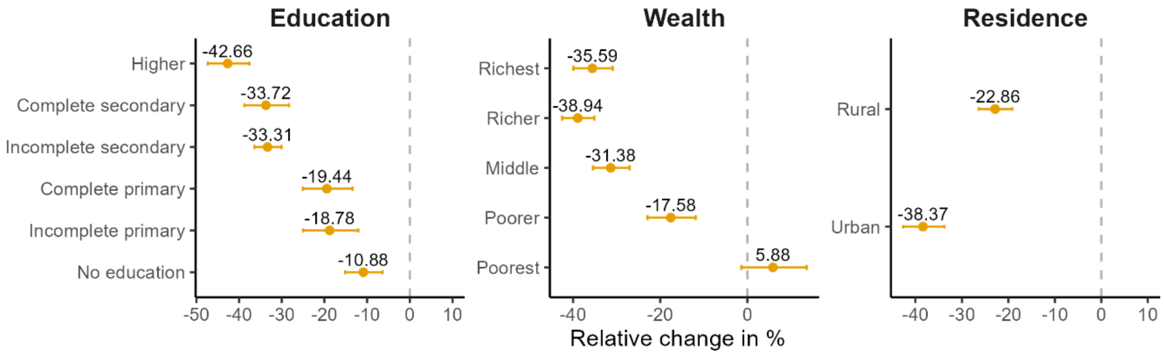


**Figure S31. Relative changes in the prevalence of adults aged 15-49 diagnosed with hypertension across subpopulations**

#### Age-adjusted

#### Tobacco consumption

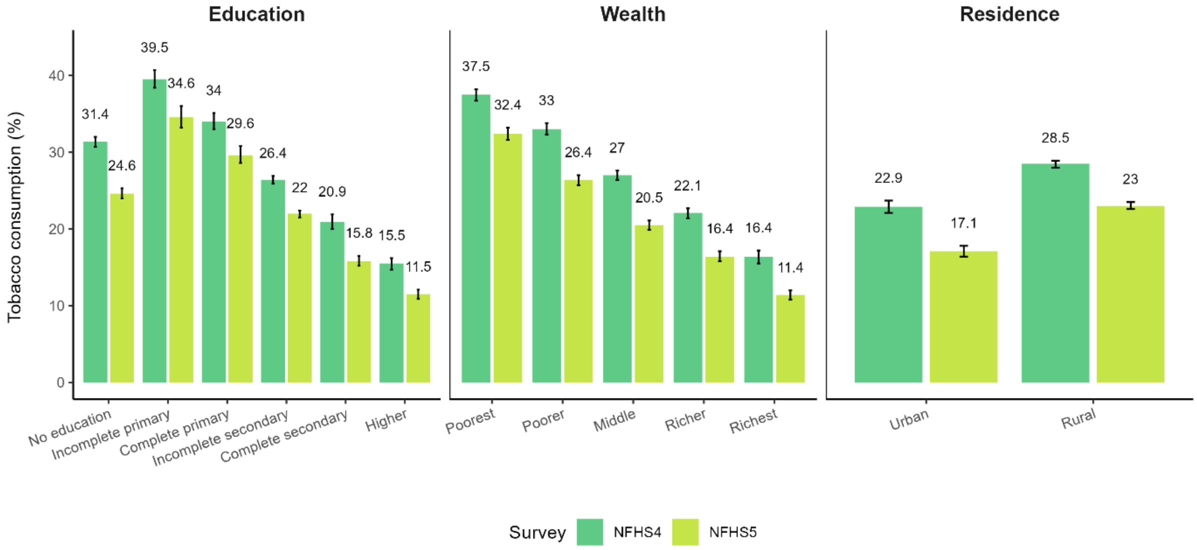


**Figure S32. Age-adjusted prevalence of tobacco consumption among adults aged 15-49 across subpopulations in each survey round**

#### Cigarette smoking

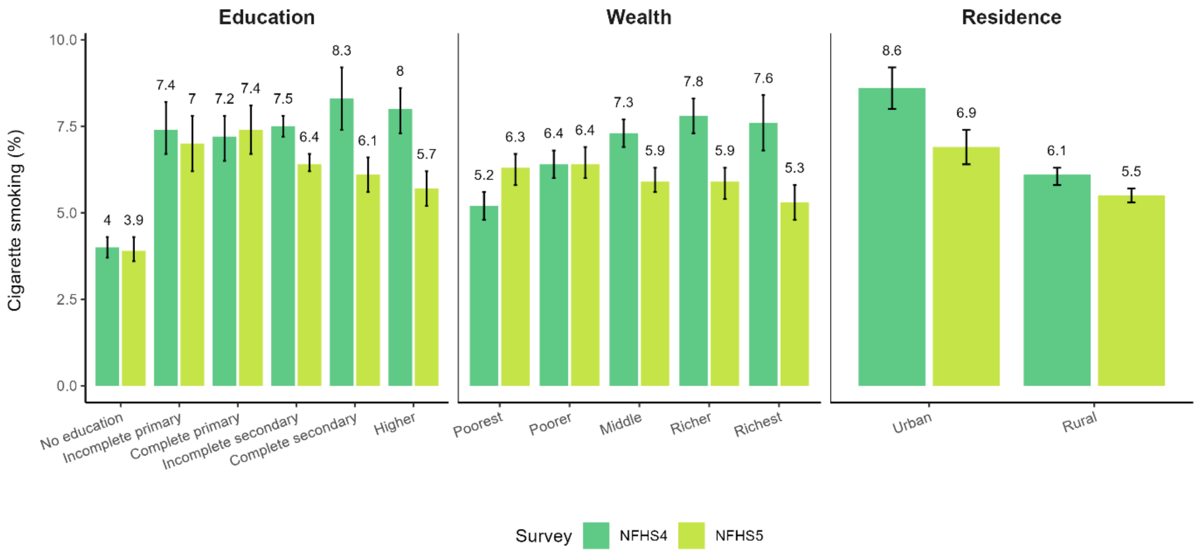


**Figure S33. Age-adjusted prevalence of cigarette smoking among adults aged 15-49 across subpopulations in each survey round**

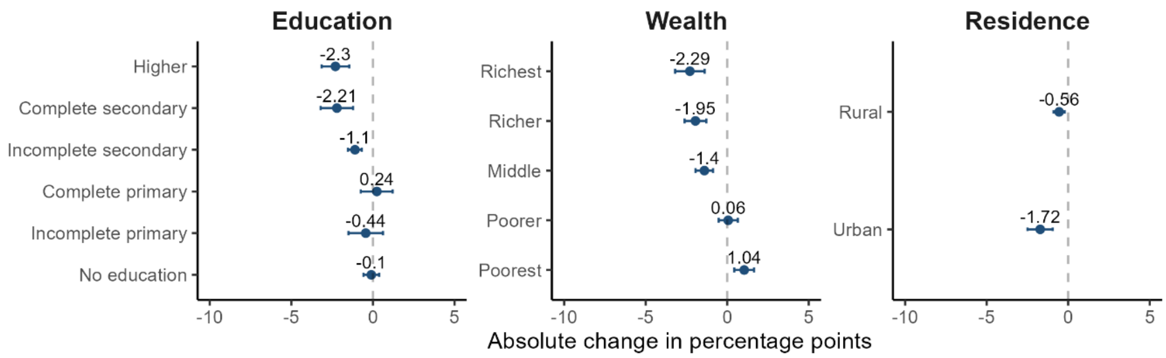


**Figure S34. Age-adjusted absolute changes in the prevalence of cigarette smoking among adults aged 15-49 across subpopulations**

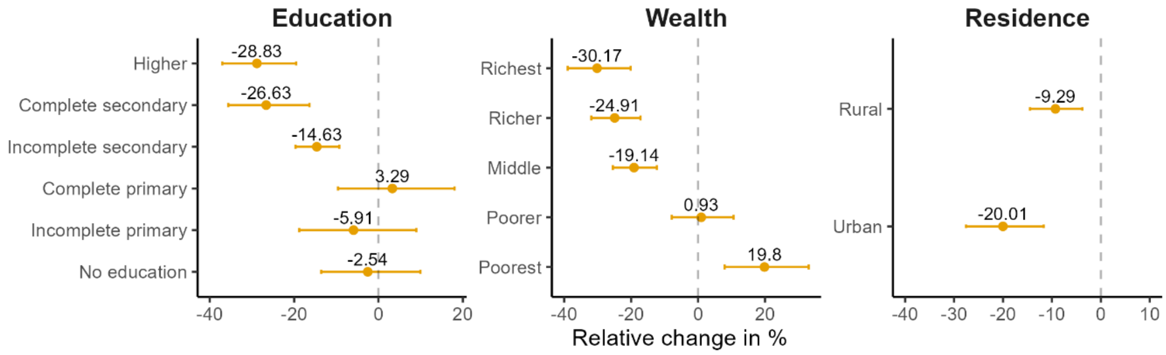


**Figure S35. Age-adjusted relative changes in the prevalence of cigarette snoking among adults aged 15-49 across subpopulations**

##### Overweight or obesity

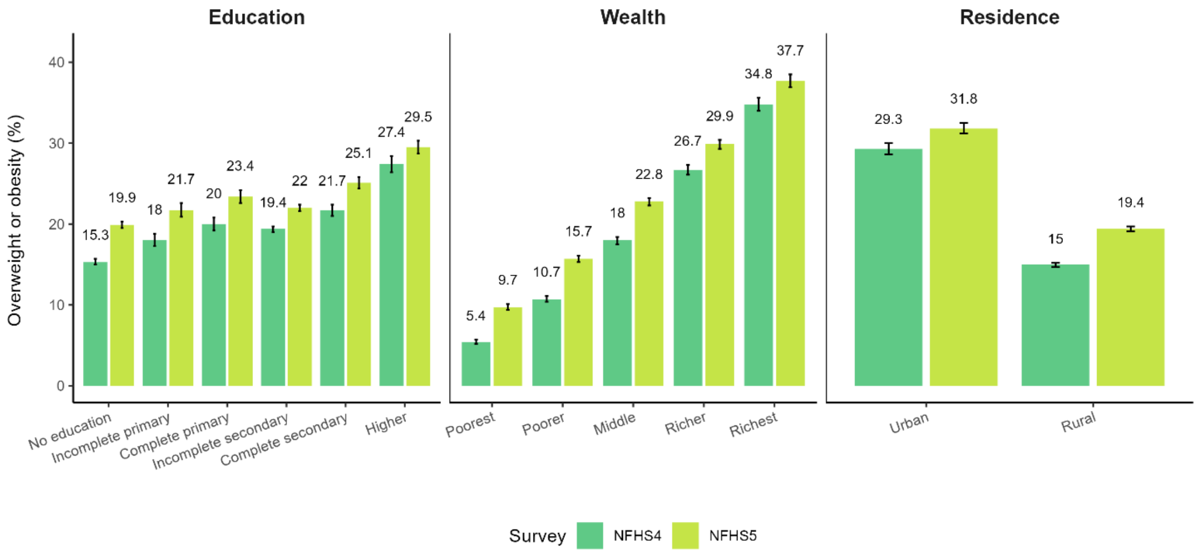


**Figure S36. Age-adjusted prevalence of overweight or obesity among adults aged 15-49 across subpopulations in each survey round**

#### Obesity

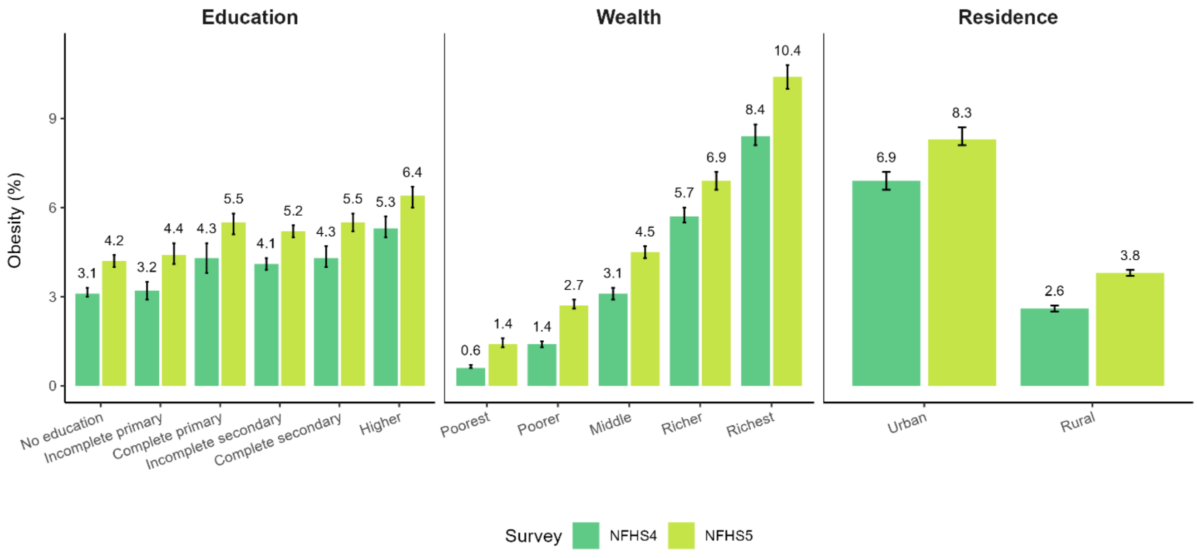


**Figure S37. Age-adjusted prevalence of obesity among adults aged 15-49 across subpopulations in each survey round**

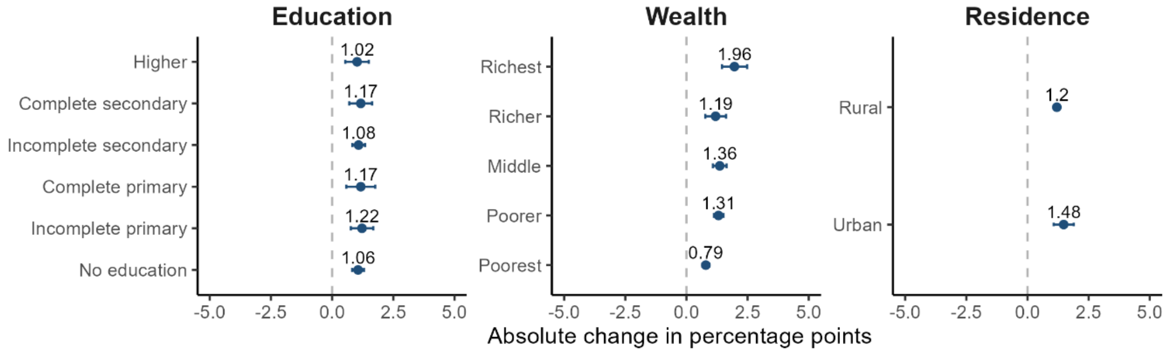


**Figure S38. Age-adjusted absolute changes in the prevalence of obesity among adults aged 15-49 across subpopulations**

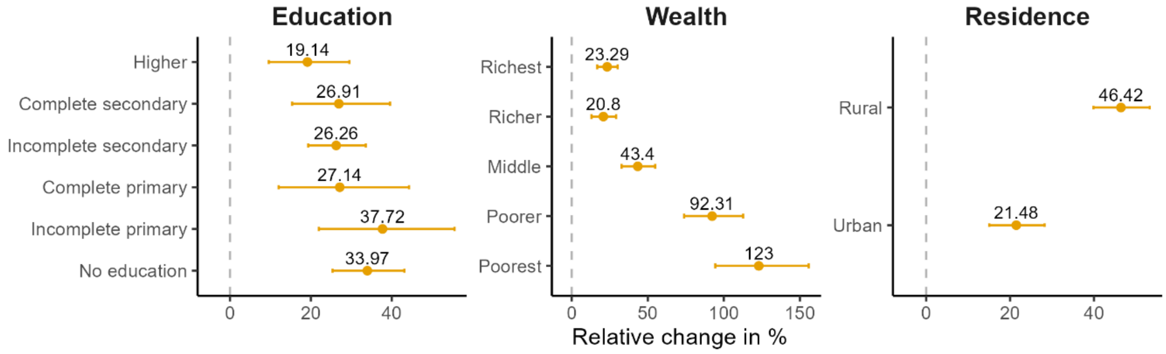


**Figure S39. Age-adjusted relative changes in the prevalence of obesity among adults aged 15-49 across subpopulations**

#### Diabetes

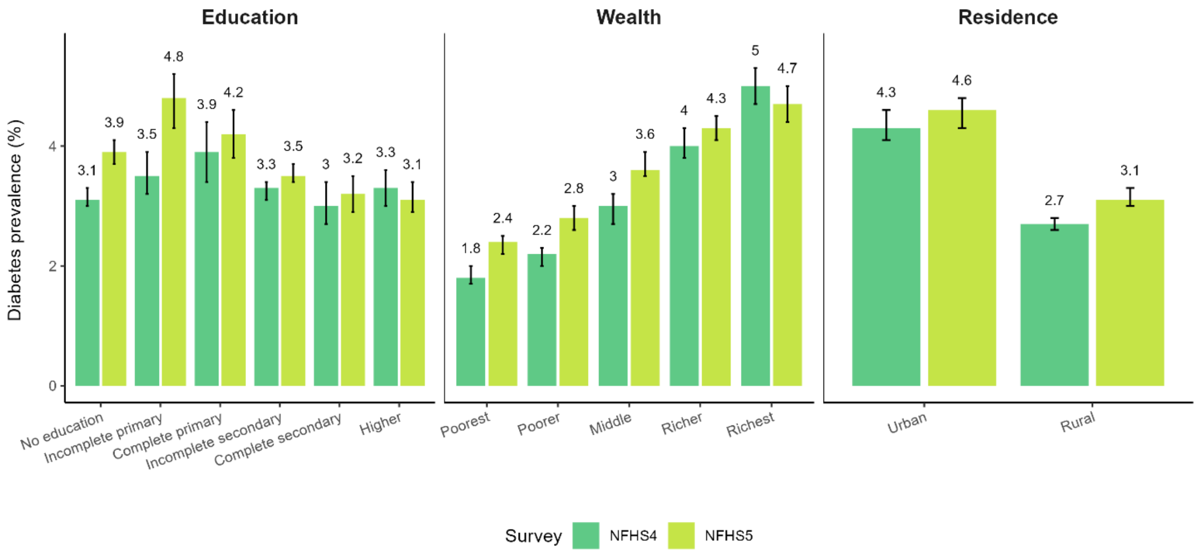


**Figure S40. Age-adjusted prevalence of diabetes among adults aged 15-49 across subpopulations in each survey round**

#### High blood glucose

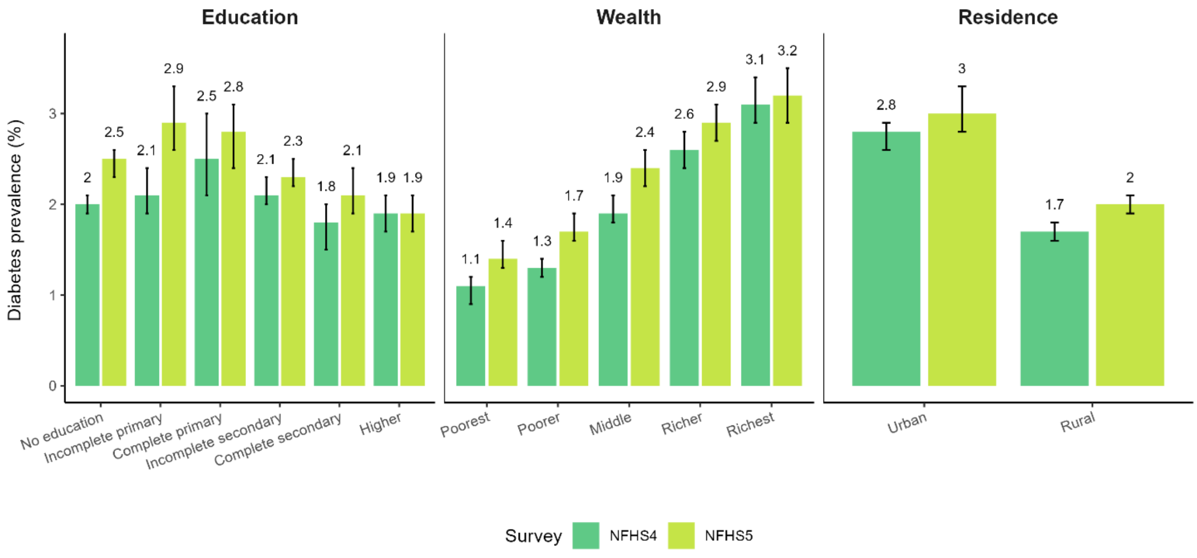


**Figure S41. Age-adjusted prevalence of high blood glucose among adults aged 15-49 across subpopulations in each survey round**

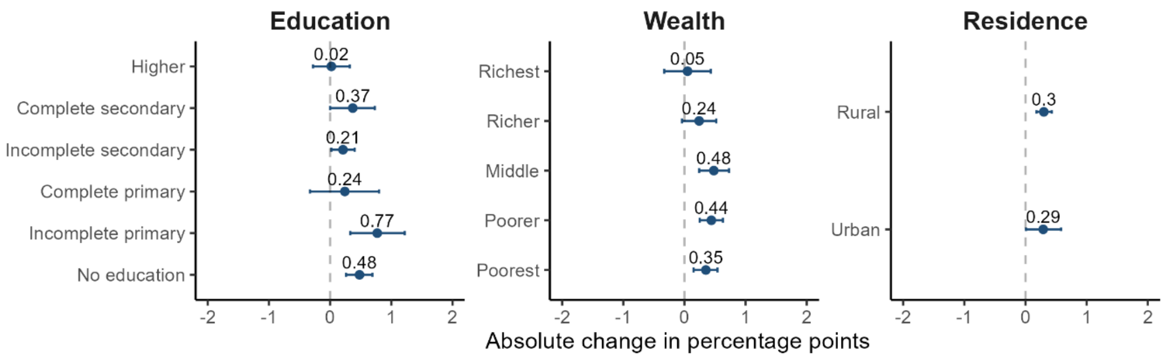


**Figure S42. Age-adjusted absolute changes in the prevalence of high blood glucose among adults aged 15-49 across subpopulations**

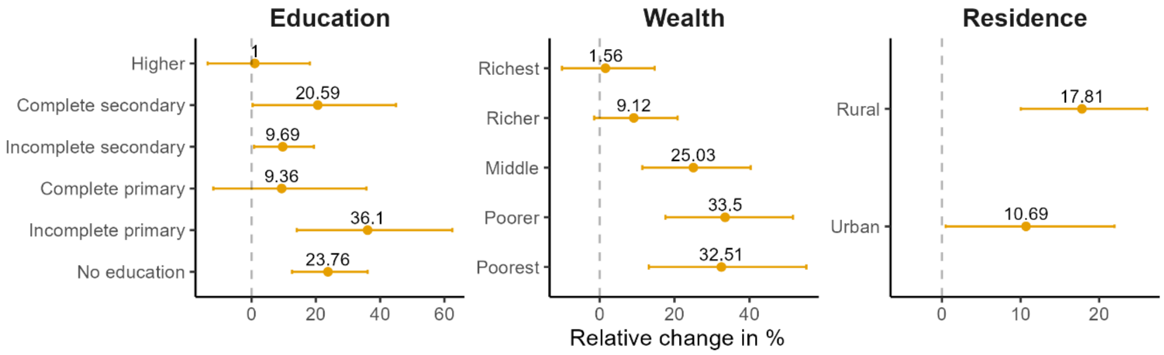


**Figure S43. Age-adjusted relative changes in the prevalence of high blood glucose among adults aged 15-49 across subpopulations**

#### Self-reported diabetes

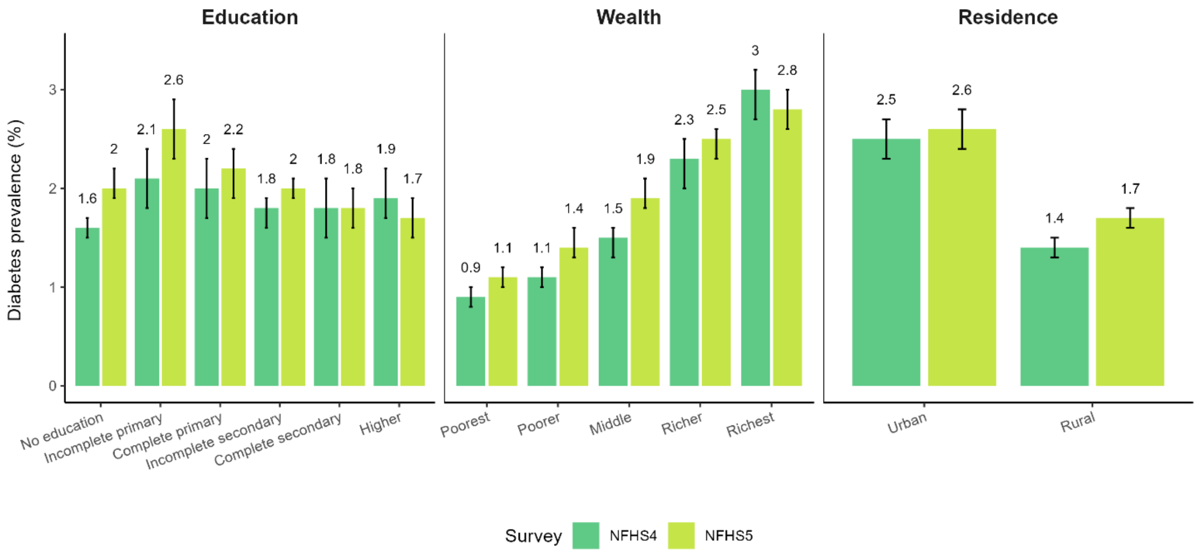


**Figure S44. Age-adjusted prevalence of self-reported diabetes among adults aged 15-49 across subpopulations in each survey round**

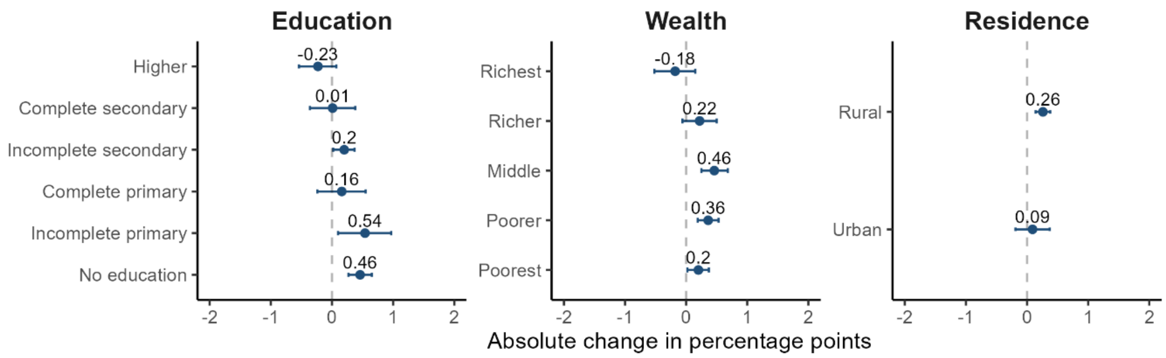


**Figure S45. Age-adjusted absolute changes in the prevalence of self-reported diabetes among adults aged 15-49 across subpopulations**

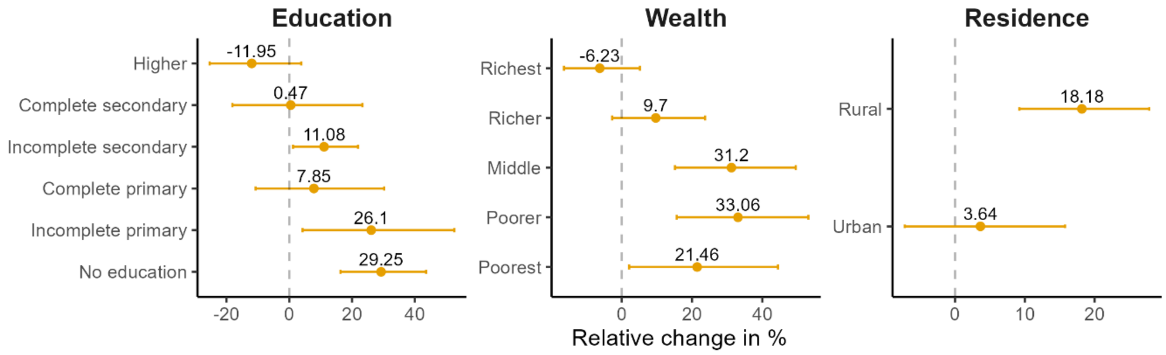


**Figure S46. Age-adjusted relative changes in the prevalence of self-reported diabetes among adults aged 15-49 across subpopulations**

#### Hypertension

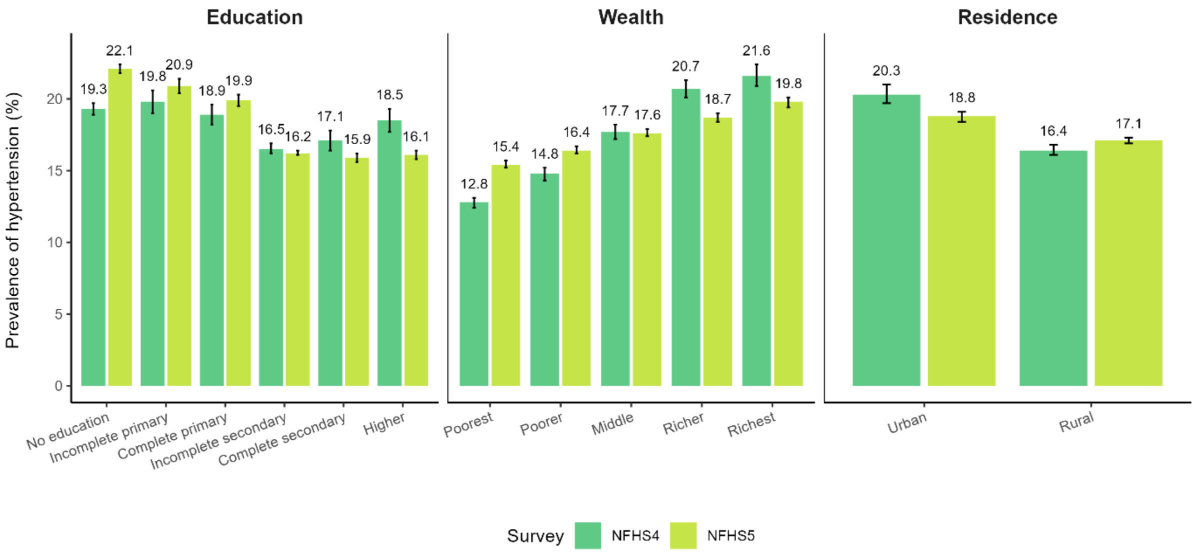


**Figure S47. Age-adjusted prevalence of hypertension among adults aged 15-49 across subpopulations in each survey round**

#### High blood pressure

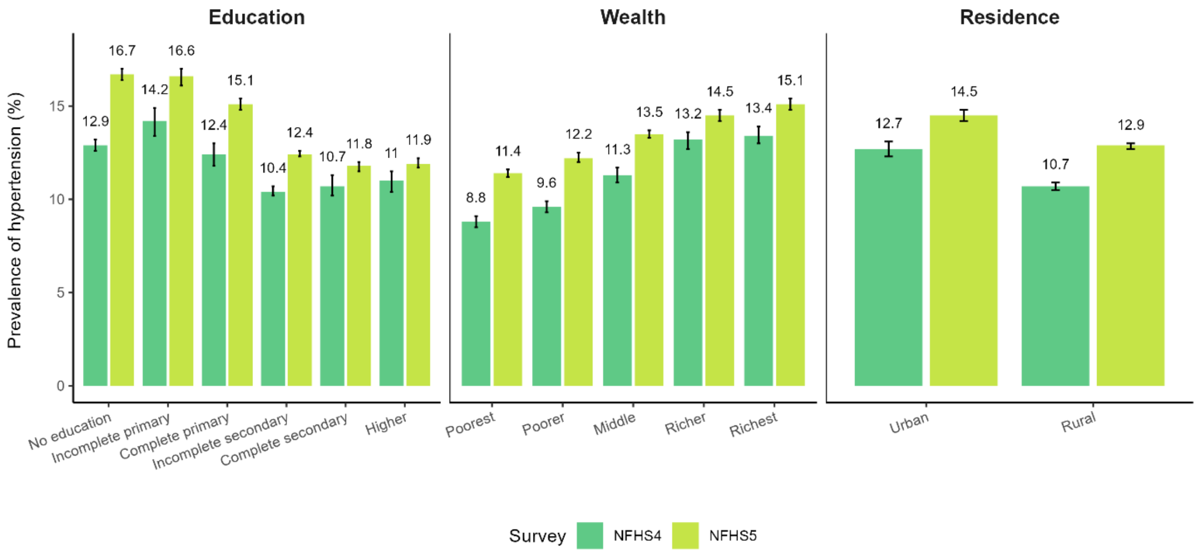


**Figure S48. Age-adjusted prevalence of high blood pressure among adults aged 15-49 across subpopulations in each survey round**

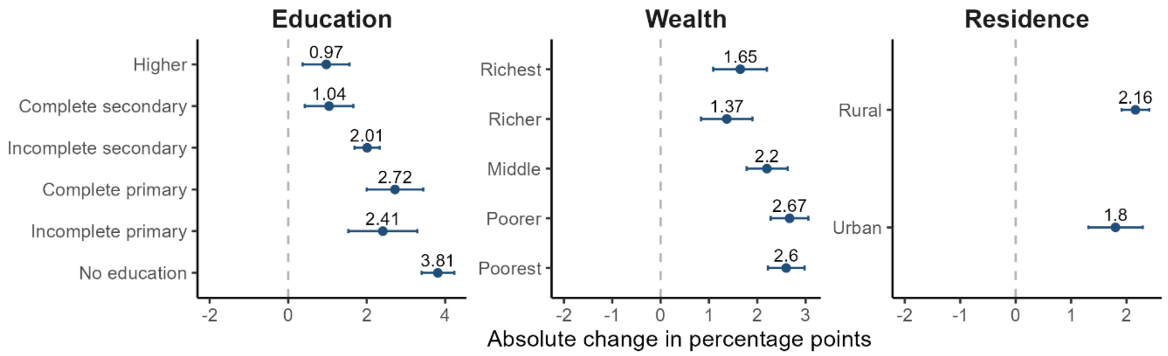


**Figure S49. Age-adjusted absolute changes in the prevalence of high blood pressure among adults aged 15-49 across subpopulations**

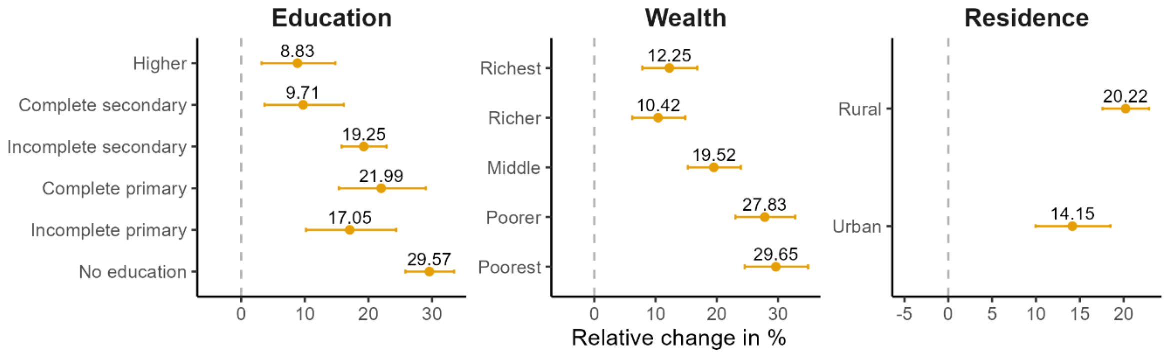


**Figure 50. Age-adjusted relative changes in the prevalence of high blood pressure among adults aged 15-49 across subpopulations**

#### Told to have high blood pressure

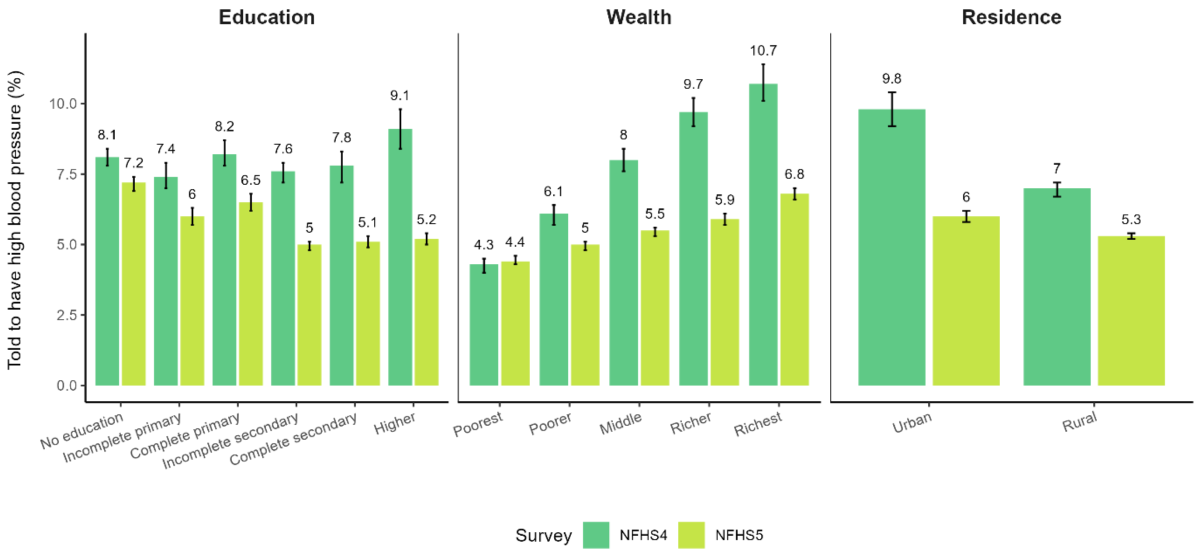


**Figure S51. Age-adjusted prevalence of adults aged 15-49 diagnosed with hypertension across subpopulations in each survey round**

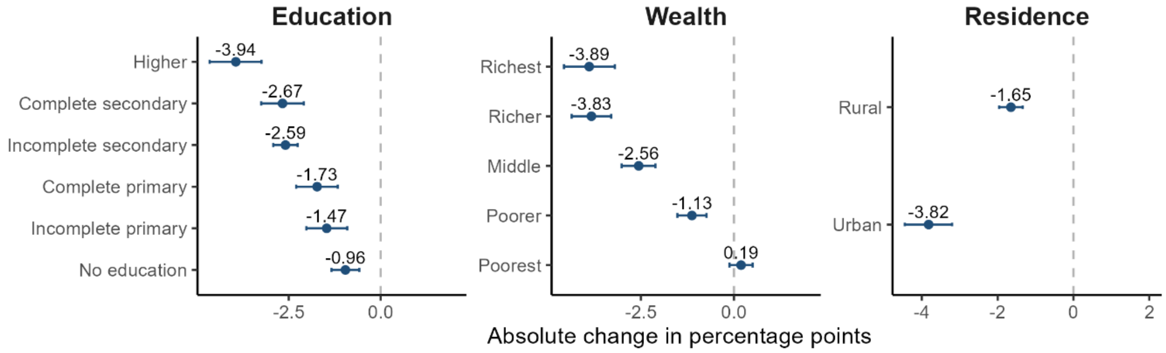


**Figure S52. Age-adjusted absolute changes in the prevalence of adults aged 15-49 diagnosed with hypertension across subpopulations**

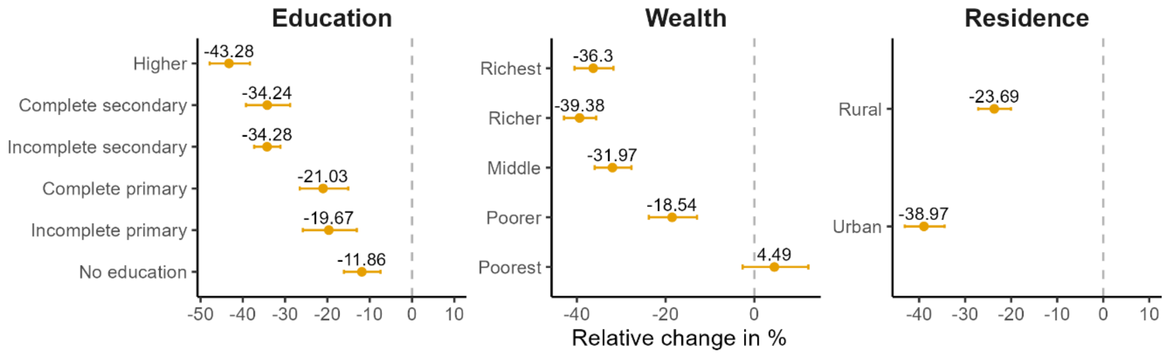


**Figure S53. Age-adjusted relative changes in the prevalence of adults aged 15-49 diagnosed with hypertension across subpopulations**

#### Results stratified by region
#### NFHS-4

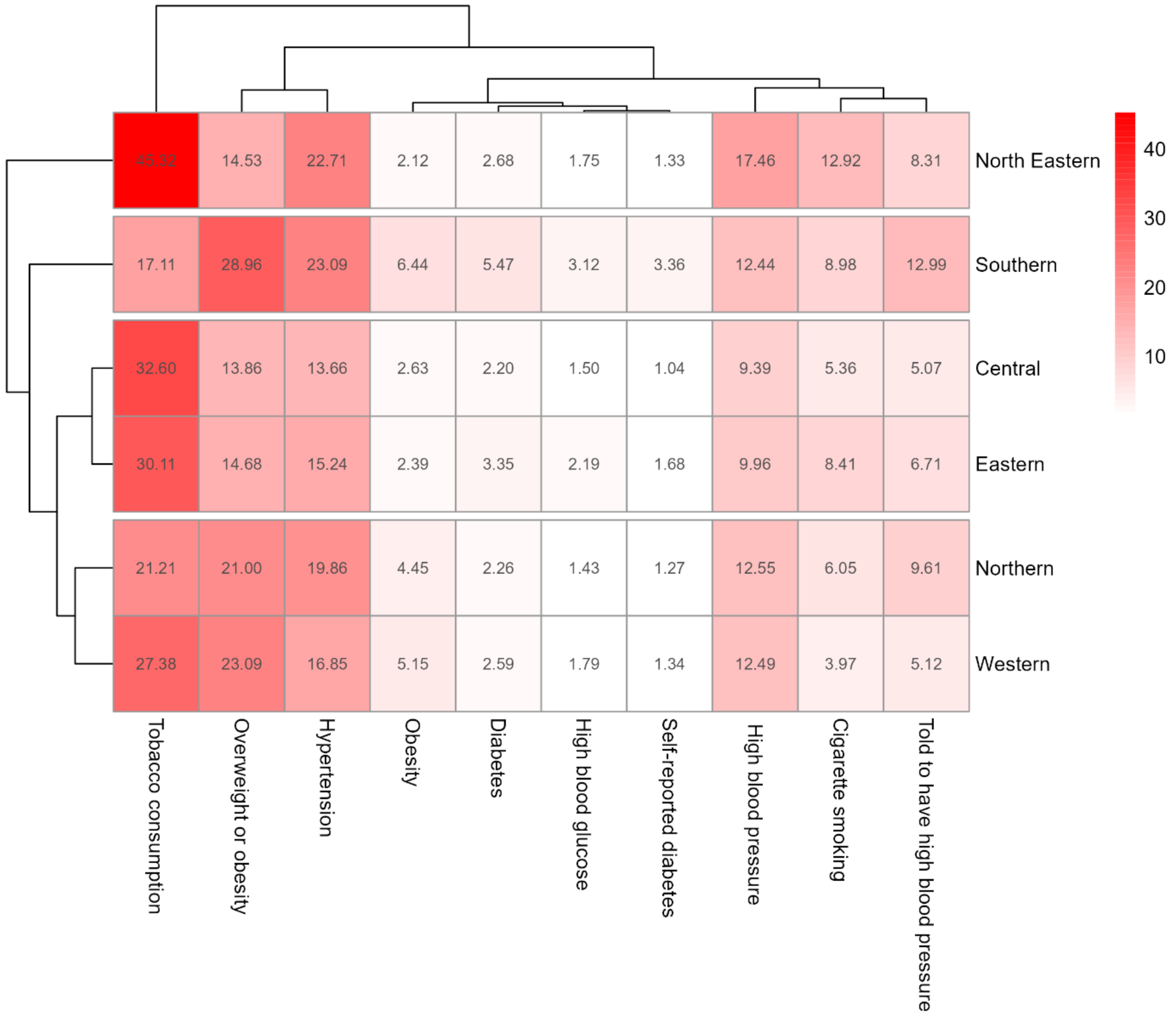


**Figure S54. Age-adjusted prevalence for each CVD risk factor across regions at the time of the NFHS-4**
The heatmap is based on hierarchical cluster analysis with average linkage and Euclidean distance.

#### Absolute changes

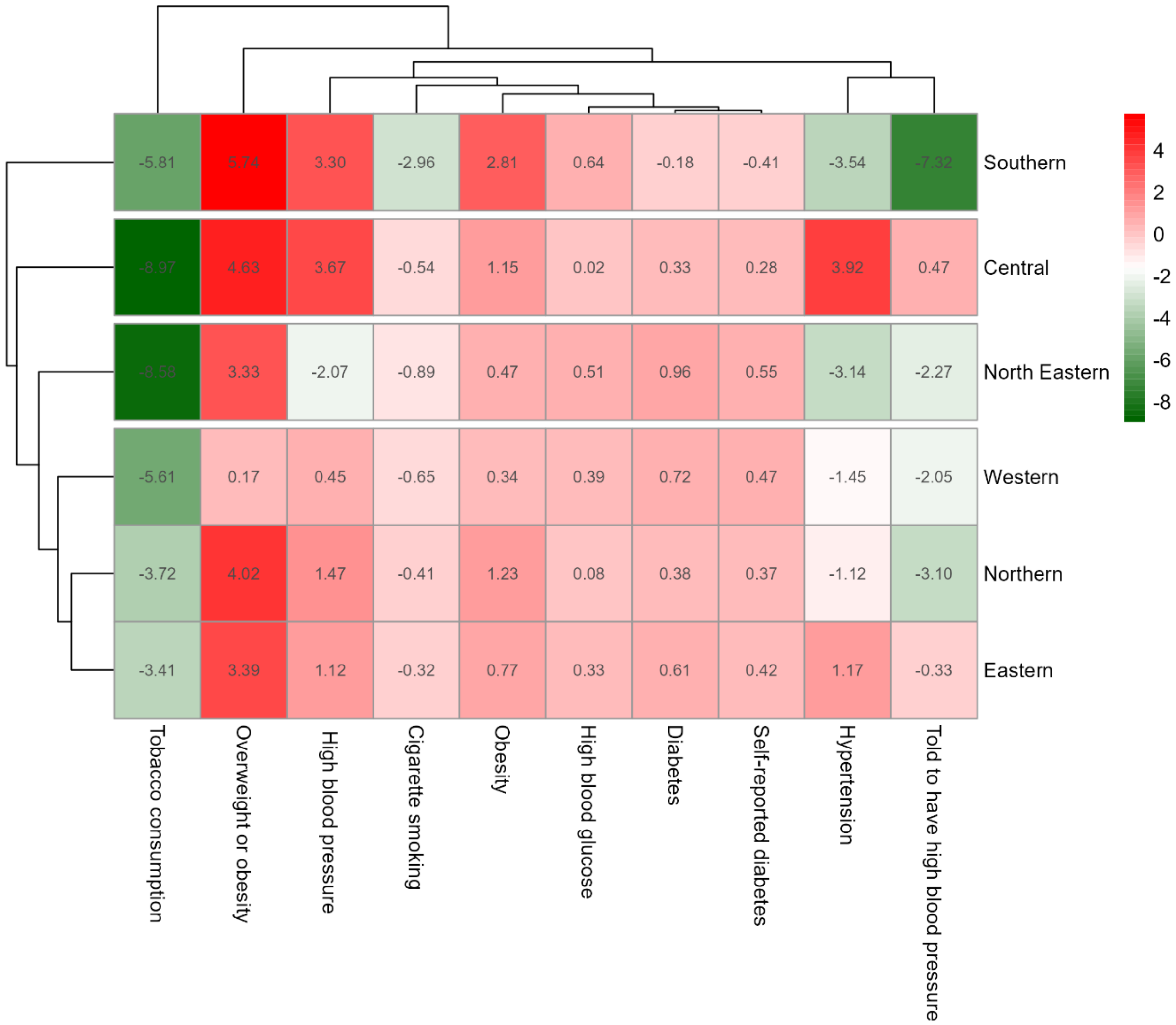


**Figure S55. Age-adjusted absolute change in the prevalence of each CVD risk factor across regions between the NFHS-4 and the NFHS-5**

The heatmap is based on hierarchical cluster analysis with average linkage and Euclidean distance.

#### Relative changes

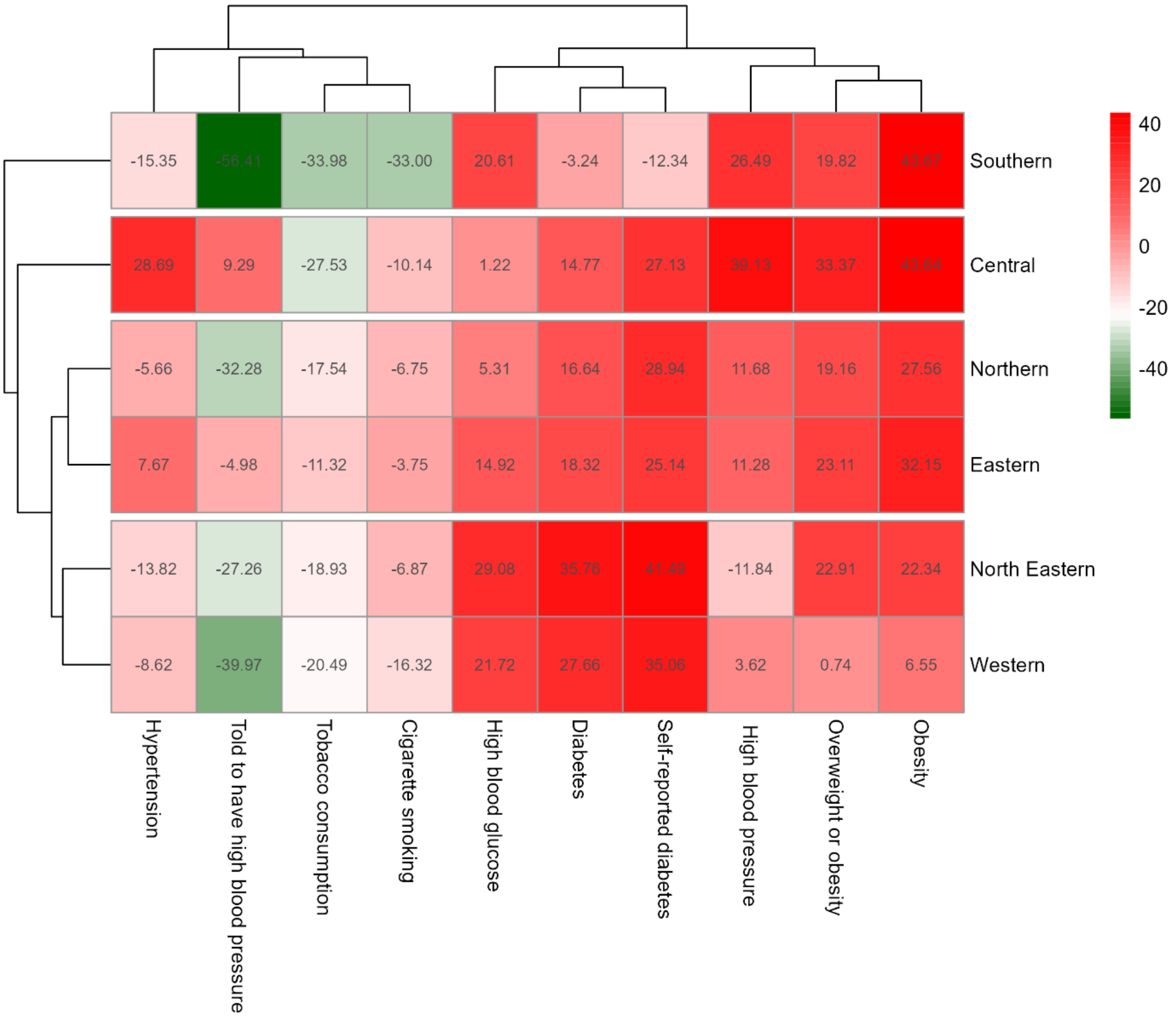


**Figure S56. Age-adjusted relative change in the prevalence of each CVD risk factor across regions between the NFHS-4 and the NFHS-5**

The heatmap is based on hierarchical cluster analysis with average linkage and Euclidean distance.

#### Results stratified by regional level of development
#### NFHS-4

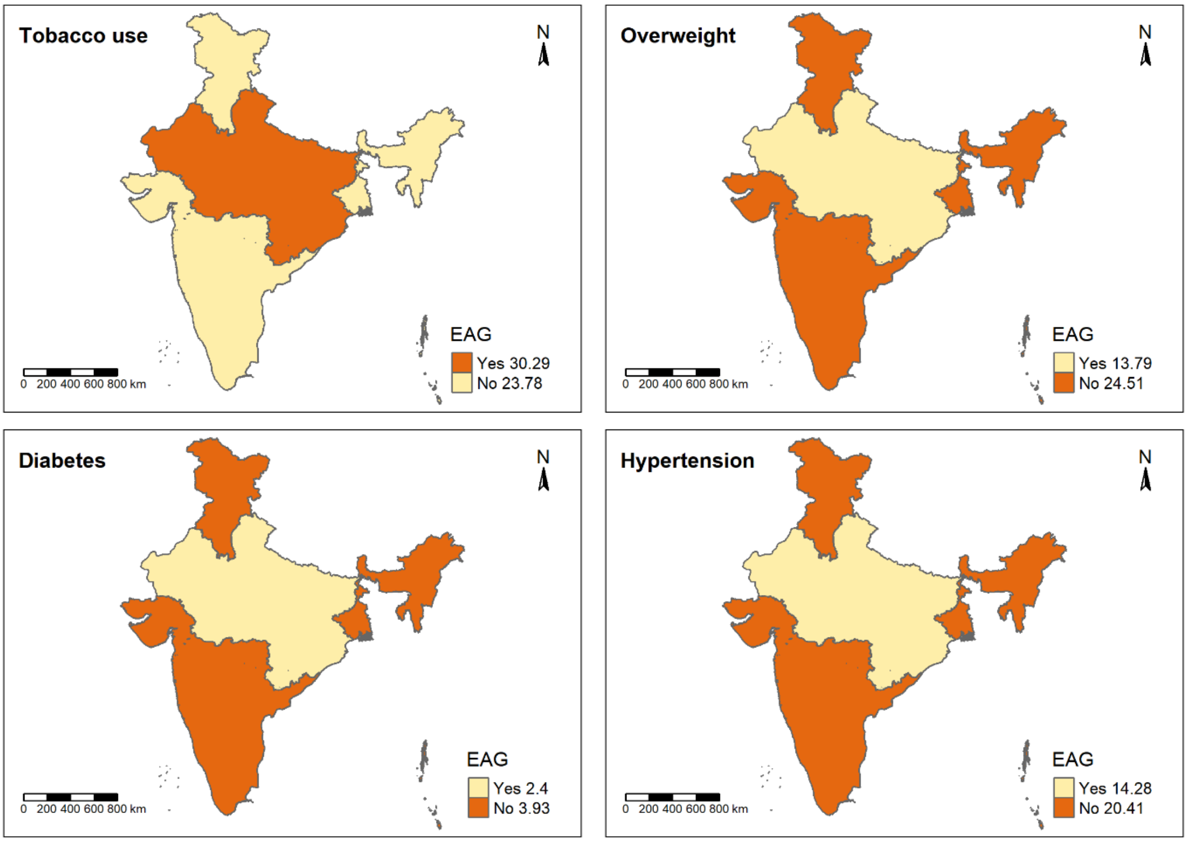


**Figure S57. Age-adjusted prevalence of CVD risk factors in the Empowered Action Group (EAG) states compared to other states and union territories at the time of the NFHS-4.**

#### Absolute changes

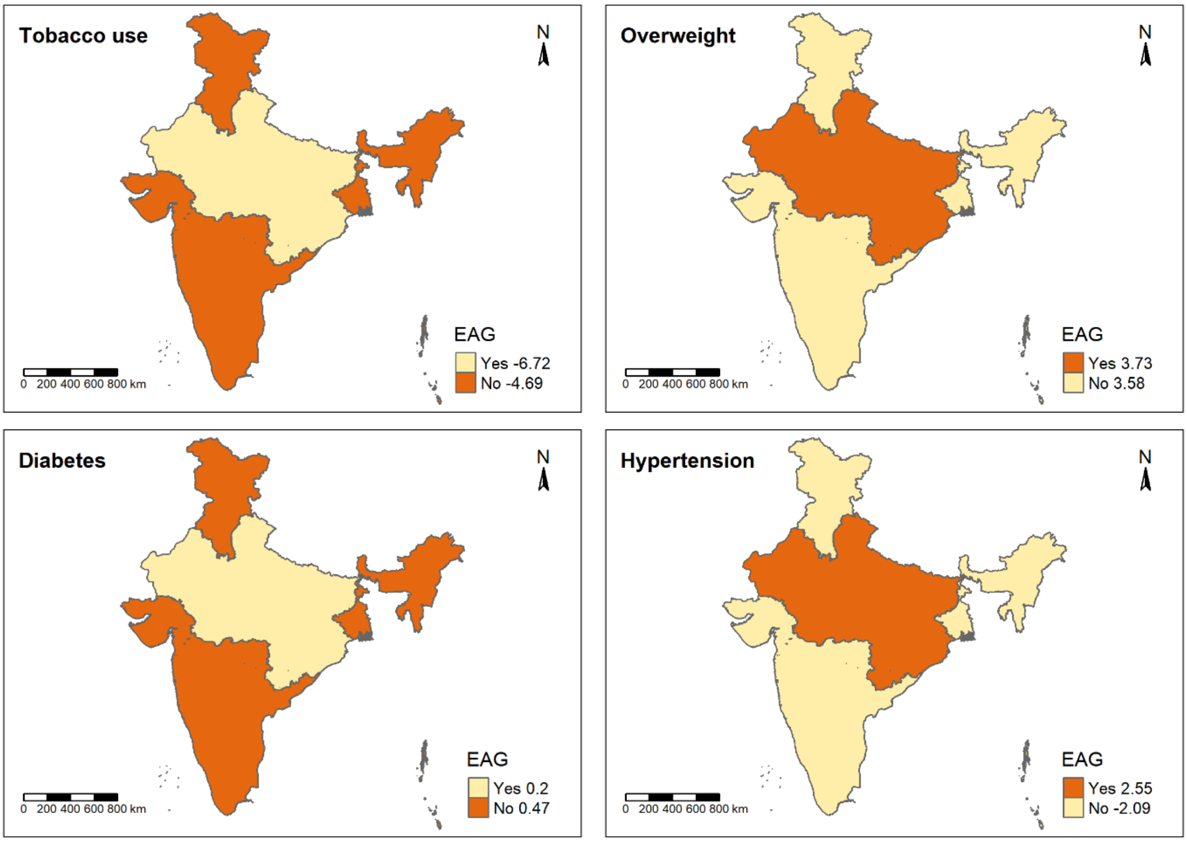


**Figure S58. Age-adjusted absolute changes (percentage points) in the prevalence of CVD risk factors in the Empowered Action Group (EAG) states compared to other states and union territories**

#### Relative changes

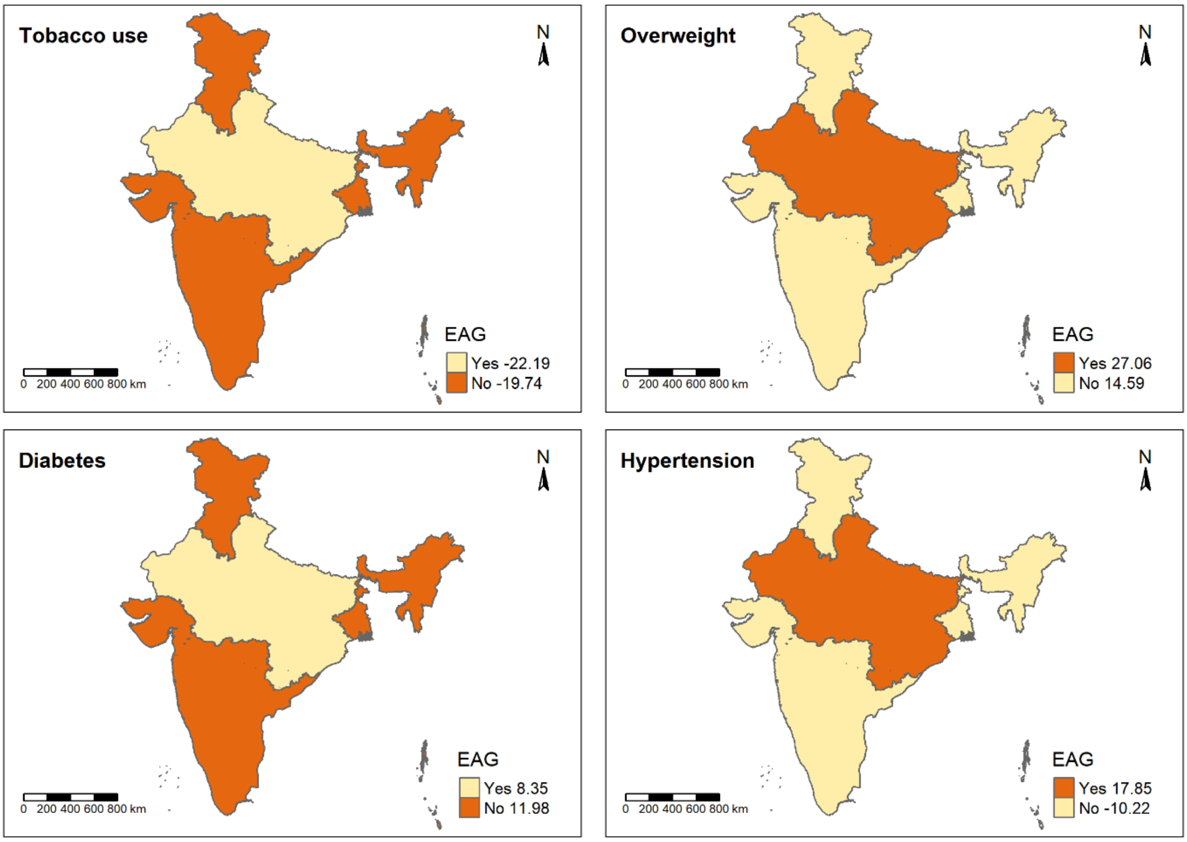


**Figure S59. Age-adjusted relative changes (%) in the prevalence of CVD risk factors in the Empowered Action Group (EAG) states compared to other states and union territories**

#### Results stratified by sex and socioeconomic status or place of residence
#### Women

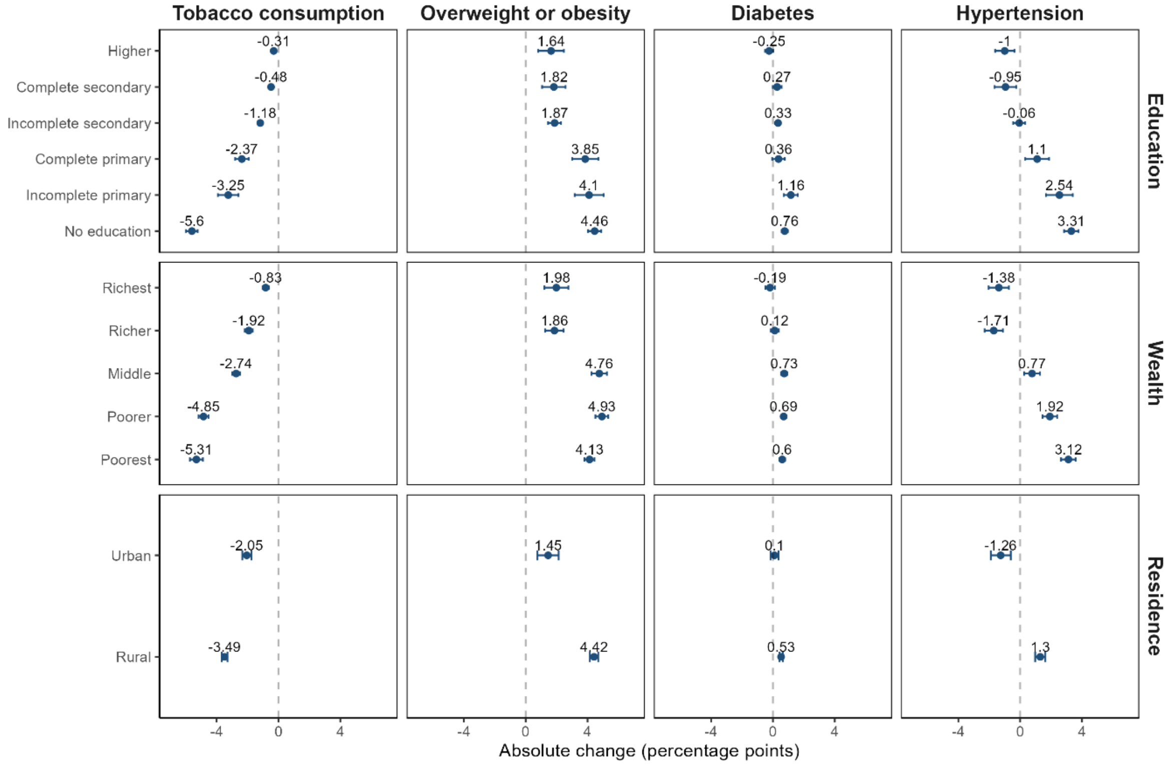


**Figure S60. Age-adjusted absolute change in each CVD risk factor among women aged 15-49 by wealth quintile, level of education, and place of residence.**

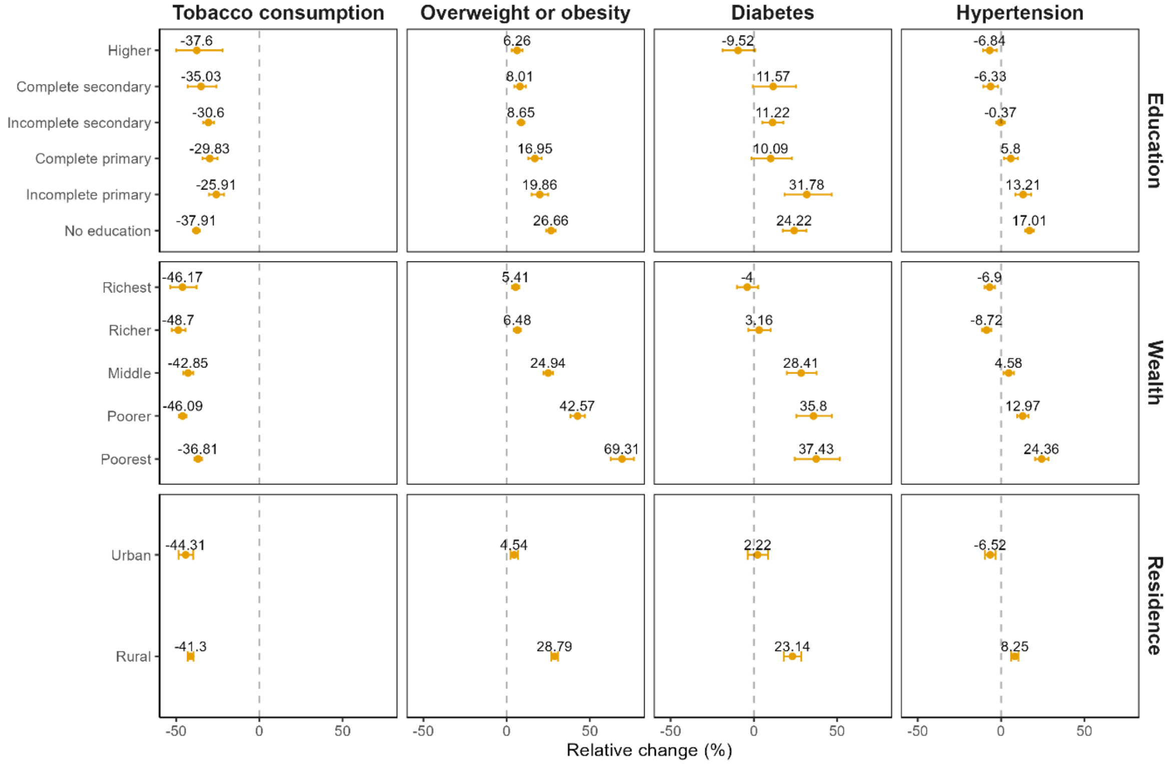


**Figure S61. Age-adjusted relative change in each CVD risk factor among women aged 15-49 by wealth quintile, level of education, and place of residence.**

#### Men

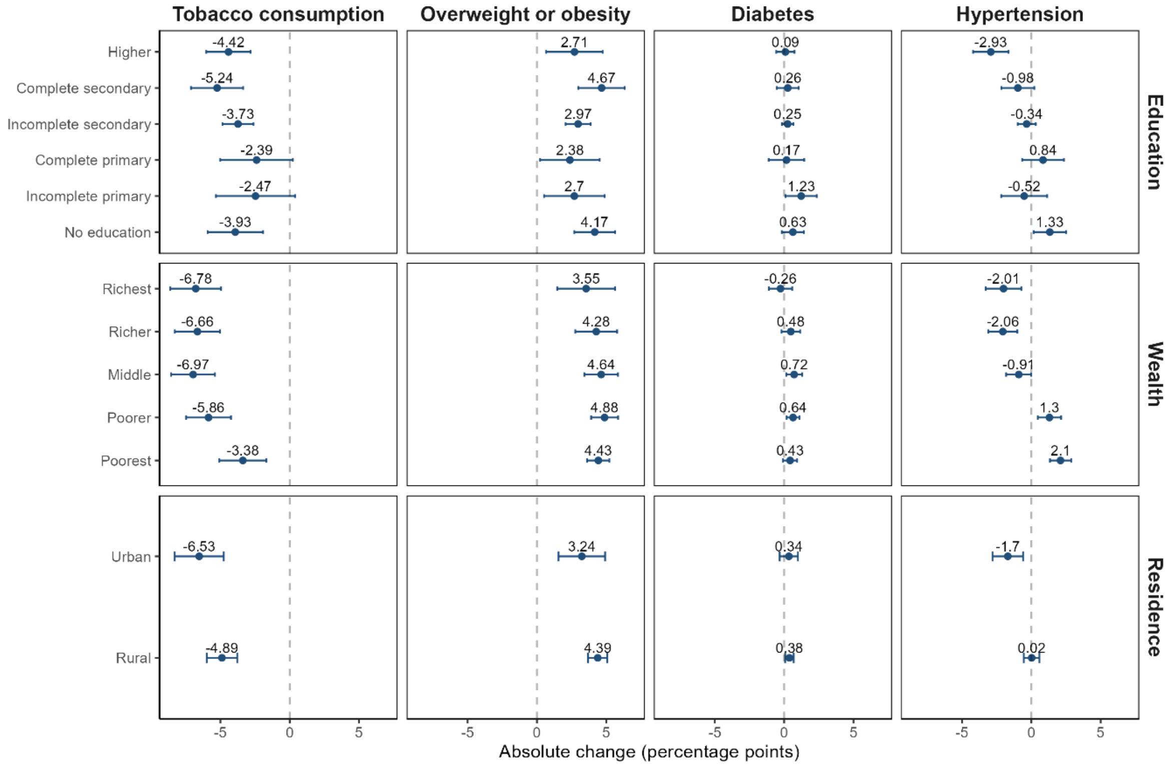


**Figure S62. Age-adjusted absolute change in each CVD risk factor among men aged 15-49 by wealth quintile, level of education, and place of residence.**

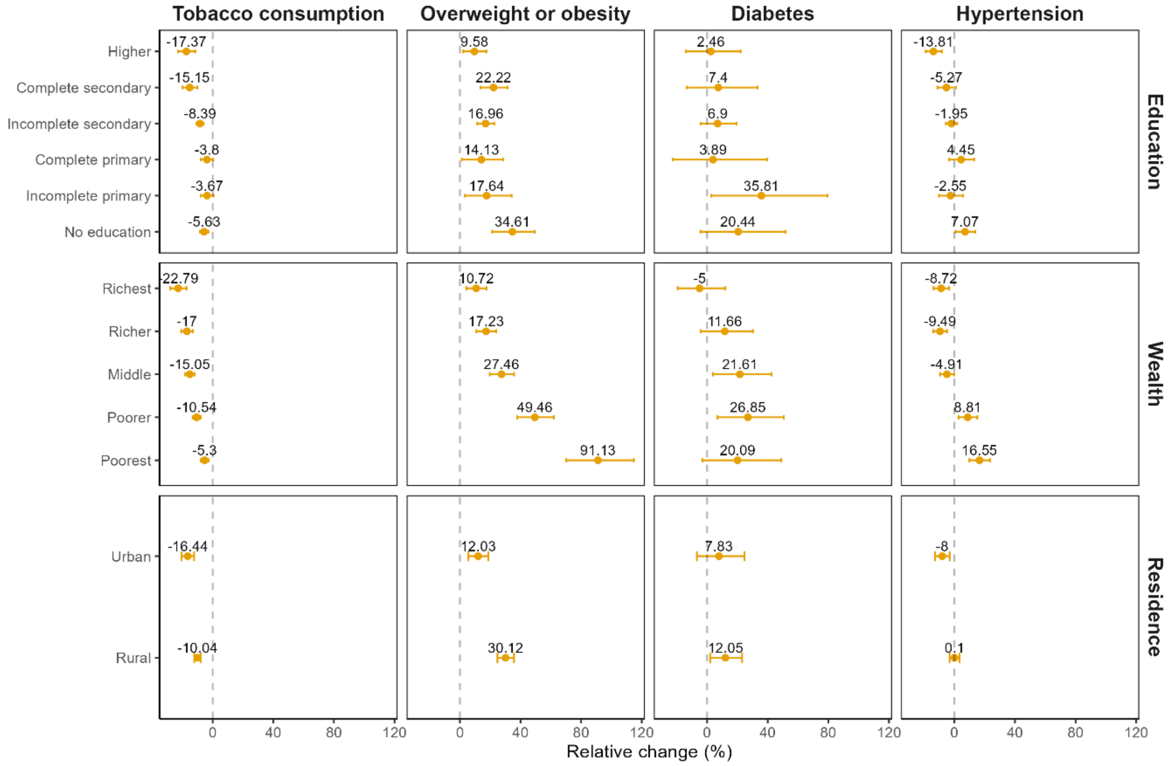


**Figure S63. Age-adjusted relative change in each CVD risk factor among men aged 15-49 by wealth quintile, level of education, and place of residence.**
